# Blood-based biomarkers and 20-year risk of clinically diagnosed Alzheimer’s disease

**DOI:** 10.64898/2026.04.16.26350847

**Authors:** Thomas J. Littlejohns, Wenyu Liu, Christopher Maronga, Tammy Y.N. Tong, Najaf Amin, Marie Breeur, Jennifer Collister, Mahboubeh Parsaeian, Keren Papier, Paolo Piazza, Gabrielle Rockett, Karl Smith-Byrne, Ruth C. Travis, Cornelia M. van Duijn, David J. Hunter

## Abstract

**Importance:** Identifying individuals in the preclinical stages of Alzheimer’s disease is necessary for inclusion into prevention trials and for early diagnosis for emerging interventions. However, this is challenging as Alzheimer’s disease pathology typically occurs in the brain at least 20 years before diagnosis.

**Objective:** To determine whether blood-based biomarkers are associated with risk of clinically diagnosed Alzheimer’s disease over a 25-year total follow-up period.

**Design:** A nested case-control study.

**Setting:** The population-based EPIC-Oxford cohort which recruited adults throughout the United Kingdom between 1993-2001.

**Participants:** 215 participants who developed incident Alzheimer’s disease were 1:1 matched with controls free of incident dementia on age at baseline, sex, and date of blood sample collection.

**Exposure:** 130 protein neuro-and inflammation-related biomarkers plus three derived ratios of biomarkers were measured from 30ml non-fasting blood samples collected at baseline assessment using the NUcleic acid-Linked Immuno-Sandwich Central Nervous System panel.

**Main Outcome:** Alzheimer’s disease was ascertained from electronic hospital inpatient and death records available up to December 2019.

**Results:** After exclusions, 213 cases and 213 controls remained (mean age at blood draw=62.8 years; women=70.9%). Over a median follow-up of 19.4 years, higher brain-derived p-tau 217 (odds ratio (OR)=1.90, 95% confidence interval (CI) 1.51, 2.40), total p-tau 217/Aβ42 (OR=1.87, 95% CI 1.47, 2.37), total p-tau 217 (OR= 1.78, 95% CI 1.43, 2.22), brain-derived p-tau 217/Aβ42 (OR=1.74, 95% CI 1.37, 2.22), brain-derived p-tau 181 (OR= 1.49, 95% CI 1.20, 1.86), total p-tau 231 (OR= 1.38, 95% CI 1.12, 1.70), and brain-derived p-tau 231 (OR= 1.37, 95% CI 1.12, 1.68) were associated with Alzheimer’s disease in models adjusted for APOE-e4. The apparent AUC for brain-derived p-tau 217 accounting for age, sex, and time of blood draw was 0.80, increasing to 0.82 with additional inclusion of APOE-e4. Additional LASSO and incremental AUC models suggested additional biomarkers did not appear to provide incremental discrimination over brain-derived p-tau 217 alone

**Conclusions and Relevance:** Blood-based biomarkers, in particular brain-derived p-tau 217, could identify individuals at-risk of Alzheimer’s disease at least two decades pre-diagnosis. This has important implications for the design of early interventions to ameliorate the pathological processes leading to Alzheimer’s disease.

**Key points:** *Question:* Do blood-based biomarkers predict risk of Alzheimer’s disease diagnosed, on average, 19 years later?

*Findings:* In 213 Alzheimer’s disease cases and 213 matched-controls, higher levels of brain-derived p-tau 181, 217 and 231, total p-tau 217 and 231, brain-derived p-tau 217/Aβ42 and total p-tau 217/Aβ42 were associated with a higher risk of Alzheimer’s disease. Brain-derived p-tau 217 was the strongest overall risk predictor, with an apparent AUC of 0.82 for Alzheimer’s disease.

*Meaning:* Blood-based biomarkers, particularly brain-derived p-tau 217, could be used to identify individuals at higher risk of Alzheimer’s disease at least two decades prior to a clinical diagnosis.

## Introduction

Substantial progress has been made in identifying modifiable risk factors for Alzheimer’s disease (AD), and in the development of disease-modifying AD therapeutics^1–3^. However, the translation of these findings into preventative trials and administration of treatments in a community setting is hampered by the long preclinical phase of AD that manifests at least 20 years prior to a diagnosis^4,5^.

Well-established diagnostic techniques, such as Positron Emission Tomography (PET) scans and cerebrospinal fluid (CSF) measures, are expensive and often inaccessible, so there is a growing shift towards the use of blood-based biomarkers for early AD detection^6^. The 2024 revised diagnosis and staging criteria for Alzheimer’s disease incorporated plasma phosphorylated-tau (p-tau) 217, 181, and 231, and amyloid beta (Aβ)-42 as core diagnostic markers of AD neuropathologic change^7^. P-tau 217, which detects Aβ plaque accumulation prior to neurofibrillary tau tangle aggregation, has emerged as the most promising blood-based biomarker for early AD detection^8–11^.

Recent assay developments have enabled the measurement of brain-derived p-tau isoforms which reflect central, rather than peripheral, nervous system pathological changes, however, the long-term risk predictive performance of these novel measures for AD have not been assessed^12,13^. Further, the majority of evidence for blood-based biomarkers has been derived from case-series studies and secondary care populations. Population-based studies have found longitudinal associations between blood-based biomarkers and AD risk, however, these have generally focused on a few select markers and have median follow-up periods less than 15 years^14–19^.

To address these gaps, we conducted a nested case-control study selecting participants from the population-based EPIC-Oxford cohort, which collected blood samples up to 25 years before AD diagnosis (median follow-up of 19.4 years). Biomarkers were measured using the NUcleic acid-Linked Immuno-Sandwich Assay (NULISAseq) Central Nervous System (CNS) panel, which detects 130 neuro-and inflammation-related proteins in the femtomolar range, including brain-derived p-tau isoforms. We investigated the long-term associations and predictive performance of multiple blood biomarkers and AD risk, and also explored lifestyle factors in association with biomarker levels^1^.

## Methods

### Population

The European Prospective Investigation into Cancer and Nutrition-Oxford (EPIC-Oxford) study is a population-based cohort of women and men aged ≥20 years old recruited throughout the UK between 1993-2001^20^. EPIC-Oxford was established to investigate dietary factors for chronic disease risk enriched for vegetarians (∼50% of participants). Participants were recruited through 1) nurses based in general practice (GP) surgeries in the English counties, Oxfordshire, Buckinghamshire and Greater Manchester; 2) postal invites distributed to members of various vegetarian, vegan and other diet-related organisations throughout the UK and via leaflets and advertisements in health food shops and dietary magazines. Participants completed a questionnaire collecting information on sociodemographic, lifestyle, and health-related factors. The study protocol was approved by a multicentre research ethics committee (Scotland A Research Ethics Committee) and all participants provided written informed consent.

### Biomarker measurement

Participants recruited through GP surgeries, and a subset of postal recruits (n=∼19,500), provided a 30ml non-fasting blood sample and underwent anthropometric and blood pressure examinations collected by an EPIC nurse or the participant’s GP. Whole blood was transported by post at ambient temperature to a laboratory in Norfolk, UK, where blood fractions (serum, plasma, red cells and buffy coat) were aliquoted into 0.5mil straws. All samples were stored in liquid nitrogen (−196 °C) until 2011 and subsequently in electric freezers (−80 °C)^21^.

The NULISA technology (Alamar Biosciences) uses a dual selection proximity ligation approach to quantify protein levels at high sensitivity and at a high signal-to-noise ratio^22^. The NULISAseq CNS panel includes proteins involved in amyloid and tau pathologies, neurodegeneration, and vascular, metabolic and inflammatory processes (**eTable 1**). Plasma samples from AD cases and controls were analysed using the NULISAseq CNS panel at the Centre for Human Genetics, University of Oxford.

50µl was extracted from stored samples and randomised across 5 96-well plates, with case-control pairs next to each other in random order. Each plate consisted of 86 samples, with 10 wells consisting of 4 negative controls (NC) to measure the background signal of each analyte and calculate detectability, 3 inter-plate controls (IPC) which are replicates of a pooled plasma control and used for sample normalisation, and 3 sample controls (SC) which are a replicate from a pooled plasma source independent from the IPCs and used to obtain intra-and inter-plate coefficient of variation. Each sample was spiked with the same level of an internal control protein to assess the uniformity of an assay run. The sequencing reads are converted into NULISA Protein Quantification (NPQ) values, a relative unit of measurement to aid statistical analyses, by dividing the value of each analyte per well by the IC, these IC-normalised values are divided by the analyte-specific median IC-normalised count for the IPC samples, and finally the data is rescaled and log_2_ transformed. The Aβ42/Aβ40 ratio was obtained by subtracting the NPQ of Aβ40 from Aβ42, and APOE4 was dichotomised into ≥10 NPQ (carrier) and <10 NPQ (non-carrier), consistent with manufacturer recommended cut-points.

### Case-control selection

Incident diagnoses and causes of death are available in EPIC-Oxford through cohort-wide linkage to electronic medical records up to December 2019. Case and control selection was restricted to participants recruited in England only, to account for country-specific variation in AD incidence due to different diagnostic practices^23^. AD cases were defined as having a hospital inpatient diagnosis or cause of death in mortality records occurring after blood sample collection using the International

Classification of Diseases (ICD)-10 codes for AD: F00 and G30. One control matched on sex, age within 12 months and date of blood sample collection within 30 days was selected per case. Controls were restricted to participants free of all-cause dementia over the follow-up period defined using ICD-10 codes F00, F01, F02, F03, and G30.

### Statistical analysis

A total of 215 AD cases and 215 matched-controls were selected. One case with less than 50% of biomarkers detectable and one case with measures which substantially deviated from the overall plate median IC reads were excluded along with their matched control, resulting in a final sample of 213 pairs.

Conditional logistic regression conditioned on age at blood draw, sex and date of blood draw was performed to obtain the odds ratio (OR) and 95% confidence intervals (CI) for the association between each continuous biomarker standardised to the control distribution and risk of AD (Model 1). The main analyses included adjustment for APOE-e4 carrier status measured on the NULISAseq CNS panel (Model 2). A secondary analysis adjusting for the following lifestyle covariates was conducted: education (degree or equivalent, higher secondary level, lower secondary level, no qualifications, unknown), smoking status (never, former, current), alcohol intake in grams per day, physical activity (very active, moderately active, low activity, inactive, unknown^24^), body mass index (BMI) per kg/m^2^, and self-reported presence of a health condition (no, yes for ≥1 of myocardial infarction, angina, stroke, high blood pressure, high cholesterol, diabetes and cancer). A missing indicator method was applied to account for 33 missing BMI values, while all other covariates had complete data. To account for multiple testing, the Benjamini-Hochberg false discovery rate (FDR) procedure was applied^25^.

All further analyses were conducted using biomarkers identified as statistically significant after FDR correction from Model 2. Biomarkers were categorised into fifths based on the distribution amongst the controls, and conditional logistic regression was performed to determine the association with AD with the middle fifth as the reference group.

The area under the receiver operating characteristic curve (AUC) were obtained for models including each biomarker 1) alone and 2) APOE-e4, with additional analyses performed to obtain the pair-level bootstrap optimism-corrected AUC. Secondary analyses were performed to obtain the AUC’s with inclusion of lifestyle factors in the prediction models both without and with APOE-e4. To assess whether adding an additional biomarker improves risk prediction, incremental ΔAUC was estimated by adding each biomarker singly to a base model containing the biomarker with the largest OR in association with AD from the original conditional logistic regression analyses.

LASSO with 5-fold cross-validation was performed, with the matching factors included in the logistic LASSO regression model as unpenalized variables. Folds were defined at the pair level, with cases and controls were balanced within folds. Model selection was based on lambda.1se corresponding to the most parsimonious model within one standard error of the highest mean cross-validated AUC. AUCs obtained from LASSO analysis were exploratory but not as the final prediction metrics since ordinary logistic LASSO rather than conditional logistic regression was considered.

Analyses were conducted in R (version 4.1.0), with *survival::clogit* used for the conditional logistic regression models, *glmnet* for (ordinary) logistic LASSO regression and *plotROC* for the AUC’s.

## Results

Cases and controls were 1:1 matched on age at blood-draw (mean = 63 years old) and sex (women = 71%. There were no significant differences between cases and controls for sociodemographic, lifestyle or health-related baseline characteristics (Table 1). The median follow-up time from blood draw to AD diagnosis was 19.4 years (interquartile range = 16.8-21.9 years), with the earliest and latest AD diagnoses occurring 10.2 and 24.9 years after blood draw, respectively. Boxplots displaying the biomarkers distributions by cases and controls are provided in **eFigure 1** and correlations between all biomarkers in **eFigure 2**.

**Table 1.** Baseline characteristics of Alzheimer’s disease cases and matched controls.

| Characteristics, N (%) | Total sample<br>(N=426) | AD Cases<br>(N=213) | Controls<br>(N=213) |
| --- | --- | --- | --- |
| Age at blood draw, mean (SD) | 62.79 (6.16) | 62.80 (6.17) | 62.77 (6.17) |
| Women | 302 (70.9%) | 151 (70.9%) | 151 (70.9%) |
| Highest qualification |  |  |  |
| Degree or equivalent | 78 (18.3%) | 43 (20.2%) | 35 (16.4%) |
| Higher secondary level | 71 (16.7%) | 35 (16.4%) | 36 (16.9%) |
| Lower secondary level | 59 (13.8%) | 26 (12.2%) | 33 (15.5%) |
| No qualifications | 151 (35.4%) | 78 (36.6%) | 73 (34.3%) |
| Unknown | 67 (15.7%) | 31 (14.6%) | 36 (16.9%) |
| Smoking status |  |  |  |
| Current | 29 (6.8%) | 12 (5.6%) | 17 (8.0%) |
| Former | 181 (42.5%) | 92 (43.2%) | 89 (41.8%) |
| Never | 216 (50.7%) | 109 (51.2%) | 107 (50.2%) |
| Alcohol (g/day), mean (SD) | 8.04 (10.90) | 8.56 (11.63) | 7.52 (10.14) |
| Physical activity |  |  |  |
| Inactive | 146 (34.3%) | 63 (29.6%) | 83 (39.0%) |
| Low activity | 147 (34.5%) | 74 (34.7%) | 73 (34.3%) |
| Moderately active | 56 (13.1%) | 30 (14.1%) | 26 (12.2%) |
| Very active | 30 (7.0%) | 19 (8.9%) | 11 (5.2%) |
| Unknown | 47 (11.0%) | 27 (12.7%) | 20 (9.4%) |
| BMI, mean (SD) | 24.79 (4.23) | 24.74 (4.10) | 24.85 (4.37) |
| Presence of one or more health conditions | 153 (35.9%) | 79 (37.1%) | 74 (34.7%) |
| Myocardial infarction | 16 (4.0%) | 11 (5.2%) | 5 (2.3%) |
| Angina | 27 (6.7%) | 13 (6.1%) | 14 (6.6%) |
| Stroke | 7 (1.7%) | NA | 5 (2.3%) |
| High Blood Pressure | 92 (22.3%) | 50 (23.5%) | 42 (19.5%) |
| High Cholesterol | 68 (16.6%) | 30 (14.1%) | 38 (17.8%) |
| Diabetes | 10 (2.5%) | NA | 7 (3.3%) |
| Cancer | 7 (1.7%) | 6 (2.8%) | NA |

Abbreviations: BMI, Body Mass Index, N, Number of Participants, SD, Standard Deviation, NA Numbers not available due to less than 5 in a cell.

In the main model, which included adjustment for APOE-e4, elevated levels of brain-derived p-tau 217 (OR=1.90, 95% CI 1.51, 2.40), total p-tau 217/Aβ42 (OR=1.87, 95% CI 1.47, 2.37), total p-tau 217

(OR=1.78, 95% CI 1.43, 2.22), brain-derived p-tau 217/Aβ42 (OR=1.74, 95% CI 1.37, 2.22), brain-derived p-tau 181 (OR=1.49, 95% CI 1.20, 1.86), total p-tau 231 (OR=1.38, 95% CI 1.12, 1.70), and brain-derived p-tau 231 (OR=1.37, 95% CI 1.12, 1.68) were associated with an increased risk of AD (**Figure 1**, **eTable 2**). GFAP, total p-tau 181 and brain-derived MAPT, were significant in Model 1, but were not significantly associated with AD risk after further adjustment for APOE-e4. Additional adjustment for lifestyle covariates minimally affected the effect (**eTable 2**).

**Figure 1.**
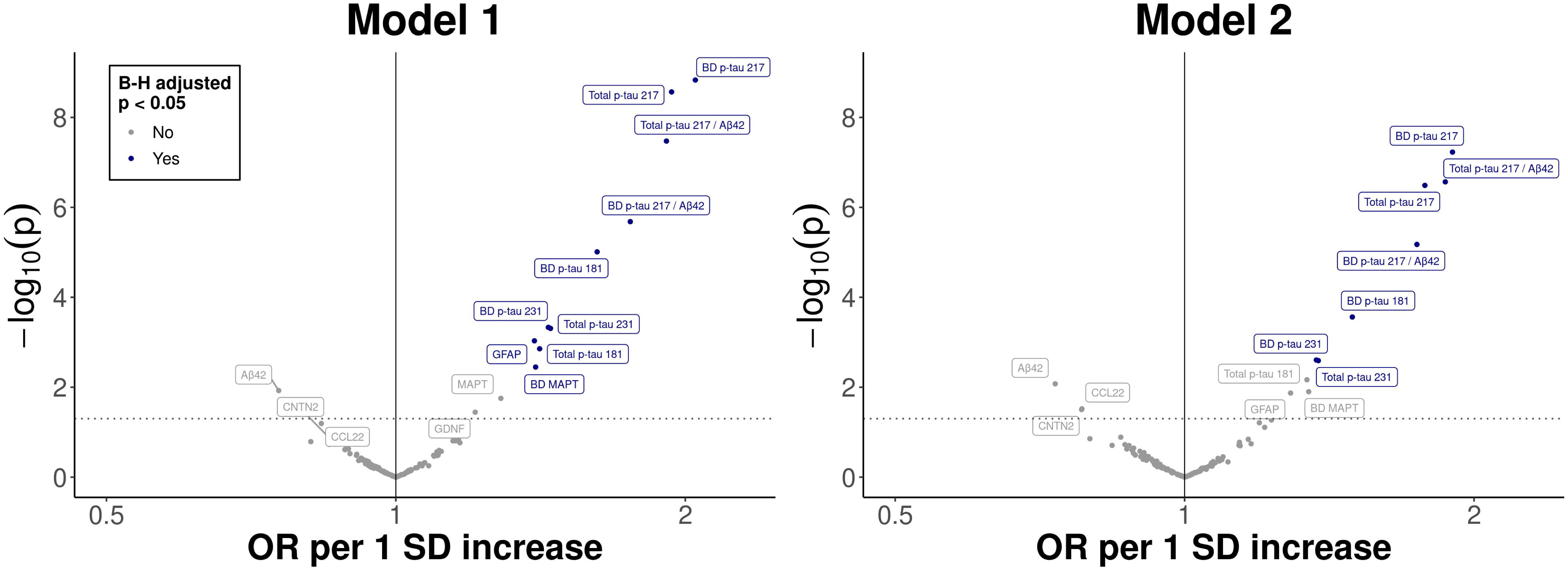
Association between blood-based biomarkers and Alzheimer’s disease risk over up to 25 years of follow-up. Conditional logistic regression models were used, accounting for matching variables including age at blood draw, sex, and date of blood draw. The points show p-values (y-axis) versus odds ratio per 1 SD increase in NPQ (standardised using the control distribution) (x-axis) for all biomarkers and risk of Alzheimer’s disease. Model 1 includes each biomarker only. Model 2 includes biomarker and APOE-e4. The horizontal grey line indicates statistical significance at p<0.05 with biomarkers labelled in blue statistically significant based on False Discovery Rate adjustment for multiple testing.

All seven biomarkers were associated with risk of AD per increase in quintile, except for total p-tau 231 after adjustment for APOE-e4 (Table 2). Brain-derived and total p-tau 217 classified the most cases in the highest quintile, 106 and 105, respectively. Compared to quintile 3, the OR’s (95% CI) for quintiles 1, 2, 4 and 5 were 0.31 (0.12, 0.79), 0.58 (0.28, 1.24), 0.71 (0.35, 1.45), and 2.77 (1.41, 5.46) for brain-derived p-tau 217, and 0.46 (0.20, 1.03), 0.69 (0.30, 1.57), 0.87 (0.43, 1.76), and 3.07 (1.51, 6.27) for total p-tau 217.

**Table 2.** Odds ratios and 95% confidence intervals for association between quintiles of blood-based biomarkers and Alzheimer’s disease.

| Biomarker, quintiles | Number of cases/controls | Biomarker only |  | Biomarker + APOE4 <sup>a</sup> |  |
| --- | --- | --- | --- | --- | --- |
|  |  | OR (95% CI) | P value for trend | OR (95% CI) | P value for trend |
| BD p-tau 181 (range) |  |  |  |  |  |
| 1 (9.44, 12.72) | 22/43 | 0.69 (0.32, 1.50) |  | 0.86 (0.38, 1.95) |  |
| 2 (12.73, 12.96) | 24/42 | 0.78 (0.38, 1.59) |  | 0.94 (0.45, 1.98) |  |
| 3 (12.97, 13.24) | 33/43 | 1 (Reference) |  | 1 (Reference) |  |
| 4 (13.24, 13.59) | 43/42 | 1.43 (0.72, 2.82) |  | 1.36 (0.67, 2.76) |  |
| 5 (13.60, 16.42) | 91/43 | 2.90 (1.51, 5.57) | <0.001 | 2.87 (1.46, 5.64) | <0.001 |
| BD p-tau 217 (range) |  |  |  |  |  |
| 1 (10.68, 11.13) | 14/43 | 0.26 (0.10, 0.67) |  | 0.31 (0.12, 0.79) |  |
| 2 (11.13, 11.24) | 26/42 | 0.54 (0.26, 1.13) |  | 0.58 (0.28, 1.24) |  |
| 3 (11.24, 11.35) | 37/43 | 1 (Reference) |  | 1 (Reference) |  |
| 4 (11.35, 11.48) | 30/42 | 0.79 (0.39, 1.57) |  | 0.71 (0.35, 1.45) |  |
| 5 (11.48, 13.70) | 106/43 | 3.26 (1.69, 6.28) | <0.001 | 2.77 (1.41, 5.46) | <0.001 |
| BD p-tau 231 (range) |  |  |  |  |  |
| 1 (12.53, 12.93) | 29/43 | 0.62 (0.32, 1.20) |  | 0.67 (0.33, 1.35) |  |
| 2 (12.94, 13.01) | 29/42 | 0.64 (0.33, 1.24) |  | 0.67 (0.34, 1.34) |  |
| 3 (13.02, 13.07) | 42/43 | 1 (Reference) |  | 1 (Reference) |  |
| 4 (13.07, 13.16) | 43/42 | 1.07 (0.60, 1.92) |  | 1.01 (0.54, 1.86) |  |
| 5 (13.17, 14.43) | 70/43 | 1.70 (0.95, 3.02) | 0.022 | 1.58 (0.86, 2.87) | 0.079 |
| Total p-tau 217 (range) |  |  |  |  |  |
| 1 (8.13, 10.06) | 16/43 | 0.44 (0.20, 0.97) |  | 0.46 (0.20, 1.03) |  |
| 2 (10.06, 10.32) | 25/42 | 0.71 (0.32, 1.58) |  | 0.69 (0.30, 1.57) |  |
| 3 (10.32, 10.57) | 33/43 | 1 (Reference) |  | 1 (Reference) |  |
| 4 (10.58, 10.82) | 34/42 | 1.06 (0.54, 2.08) |  | 0.87 (0.43, 1.76) |  |
| 5 (10.83, 14.06) | 105/43 | 3.97 (2.01, 7.84) | <0.001 | 3.07 (1.51, 6.27) | <0.001 |
| Total p-tau 231 (range) |  |  |  |  |  |
| 1 (7.95, 12.13) | 15/43 | 0.40 (0.18, 0.87) |  | 0.42 (0.19, 0.96) |  |
| 2 (12.13, 12.43) | 34/42 | 1.04 (0.54, 1.99) |  | 1.09 (0.55, 2.17) |  |
| 3 (12.44, 12.70) | 36/43 | 1 (Reference) |  | 1 (Reference) |  |
| 4 (12.71, 13.04) | 51/42 | 1.39 (0.76, 2.57) |  | 1.27 (0.67, 2.42) |  |
| 5 (13.05, 16.95) | 77/43 | 2.22 (1.20, 4.11) | <0.001 | 2.02 (1.07, 3.83) | 0.002 |
| BD p-tau 217 / Aβ42 (range) |  |  |  |  |  |
| 1 (-2.20, -0.84) | 17/43 | 0.35 (0.16, 0.74) |  | 0.38 (0.17, 0.86) |  |
| 2 (-0.83, -0.36) | 31/42 | 0.79 (0.40, 1.56) |  | 0.79 (0.39, 1.61) |  |
| 3 (-0.35, 0.13) | 37/43 | 1 (Reference) |  | 1 (Reference) |  |
| 4 (0.14, 0.90) | 44/42 | 1.6 (0.79, 3.23) |  | 1.37 (0.65, 2.88) |  |
| 5 (0.92, 5.01) | 84/43 | 3.33 (1.72, 6.44) | <0.001 | 3.59 (1.79, 7.22) | <0.001 |
| Total p-tau 217 / Aβ42 (range) |  |  |  |  |  |
| 1 (-2.73, -1.72) | 15/43 | 0.39 (0.18, 0.84) |  | 0.45 (0.20, 1.01) |  |
| 2 (-1.71, -1.18) | 27/42 | 0.72 (0.36, 1.45) |  | 0.68 (0.33, 1.41) |  |
| 3 (-1.17, -0.73) | 31/43 | 1 (Reference) |  | 1 (Reference) |  |
| 4 (-0.72, 0.04) | 52/42 | 1.81 (0.92, 3.56) |  | 1.5 (0.74, 3.04) |  |
| 5 (0.05, 4.76) | 88/43 | 3.28 (1.76, 6.08) | <0.001 | 3.34 (1.75, 6.35) | <0.001 |

Abbreviations: CI, Confidence Interval, OR, Odds Ratio
<sup>a</sup> APOE-e4 is based on the dichotomisation of APOE4 protein into carriers (≥10 NPQ) and non-carriers (<10 NPQ).

The strongest risk predictor for AD was brain-derived p-tau 217 (AUC=0.800) followed by total p-tau 217 (AUC=0.778, Figure **2**, **eTable 3**). Inclusion of APOE-e4 alone resulted in an AUC of 0.685, which increased to 0.816 with additional inclusion of brain-derived p-tau 217, and 0.795 with additional inclusion of total p-tau 217 in the prediction models. The bootstrap optimism-corrected corrected AUC reduced to 0.711 and 0.703 for brain-derived and total p-tau 217 (**eTable 3**), respectively. Adding the lifestyle variables yielded larger mean optimism to be corrected, which was a much larger penalty than the gain in apparent AUCs. This indicated that adding the lifestyle variables provided no discrimination beyond models including biomarkers with and without APOE4. Bootstrap incremental ΔAUC analyses showed that, once brain-derived p-tau 217 was included, none of the other biomarkers materially improved discrimination (**eTable4**). In LASSO analysis, the lambda.1se model yielded only a small loss of discrimination (AUC 0.70 vs 0.72) while selecting a much sparser model, with brain-derived p-tau 217 retained as the only biomarker (**eFigure 3**, **eTable5**).

**Figure 2.**
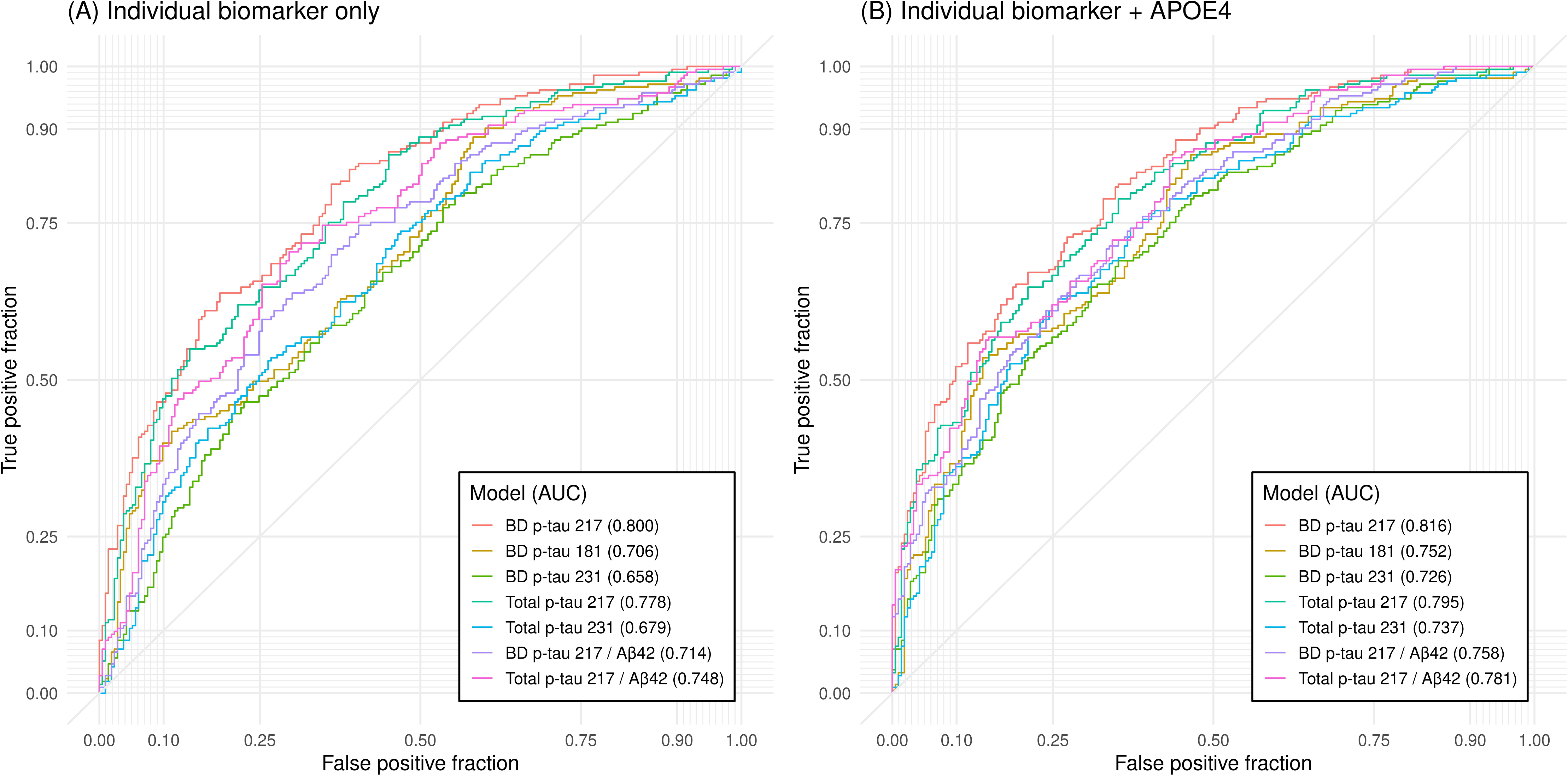
Apparent risk prediction of 25-year risk of Alzheimer’s disease for individual blood-based biomarkers (A) with individual biomarker only, (B) with individual biomarker + APOE-e4^a^. Abbreviations: AUC, Area Under the Curve, ROC, Receiver Operating Curve ^a^ APOE-e4 carrier status based on APOE4 on the NULISAseq panel The seven biomarkers were FDR-adjusted significant biomarkers based on conditional logistic regression models with biomarker + APOE4 (Model 2).

In secondary analysis restricted to controls, BMI per 5 kg/m^2^, alcohol per 10g/day intake, being an ever versus never smoking, and being physically inactive versus active were associated with higher and lower levels of various biomarkers (**eFigure 4**). However, no associations between lifestyle factors and the seven biomarkers related to AD risk were observed.

## Discussion

In this nested-case control study of participants selected from a population-based cohort, elevated brain-derived p-tau 217, total p-tau 217/Aβ42, total p-tau 217, brain-derived p-tau 217/Aβ42, brain-derived p-tau 181, total p-tau 231, and brain-derived p-tau 231 were associated with AD diagnosed clinically on average two decades after blood draw. Brain-derived p-tau 217 emerged as the strongest risk predictor for AD. When investigating the utility of multiple biomarkers for AD risk prediction, only brain-derived p-tau 217 was selected, with other markers providing no significant additional discriminative performance. These findings suggest that blood-based biomarkers are predictive of AD risk decades many years in advance of a clinical diagnosis in a population-based setting and support the utility of blood-based measures for early AD diagnosis and risk stratification.

In the current study, we used the NULISAseq CNS panel which consisted of 130 neuro-and inflammatory-proteins, and was specifically enriched for established blood-biomarkers for AD. To identify biomarkers most strongly associated with AD across the panel, we corrected for multiple comparisons. Despite the conservative nature of this analytic strategy, and accounting for the strongest risk factors for AD; age, sex, and APOE-e4; we found that all brain-derived and total p-tau isoforms, except total p-tau 181, as well as p-tau 217/Aβ-42 ratios, were associated with AD risk.

Blood-based p-tau isoforms are indicators of the early pathological hallmarks of AD, including both tau pathology in the brain as well as Aβ accumulation^26^. Evidence suggests that both these hallmarks interact to affect plasma p-tau, as blood-based p-tau levels are elevated in response to exposure to Aβ pathology but not in non-AD tauopathies^6^. Consequently, p-tau isoforms are recognised as promising markers for AD prognosis at an early pre-clinical stage^8,27,28^. Consistent with our finding for p-tau 217, a 2025 meta-analysis of 113 studies found that p-tau 217 was the highest performing p-tau isoform for identifying biologically defined AD based on Amyloid/Tau-PET, CSF markers, or neuropathology, with an AUC of 0.91, compared to p-tau 181 (AUC=0.82) and p-tau 231 (0.80)^29^.

Our findings on the associations between p-tau and AD are consistent with recent population-based longitudinal studies. In 2,148 Swedish adults, p-tau 181 and 217 were associated with a higher risk of all-cause dementia and AD over a median follow-up of 10 years, with p-tau 217 having the highest AUC (0.77) for AD risk prediction^17^. In 2 US-based populations, p-tau-181 was associated with incident dementia over a median of 7 years in 1,339 participants^19^, whilst p-tau-217 was associated with probable dementia over a median of 14 years in 2,766 women^15^. We build upon these findings, by observing these associations in blood collected a median of 19 years prior to recorded diagnosis, extending the time in which these biomarkers predict AD risk. A novel finding was that higher levels of brain-derived p-tau-181,-217, and-231 were all associated with long-term AD risk with brain-derived p-tau 217 emerging as the strongest overall predictor. There is limited research on brain-derived p-tau for AD prognosis as the assays to measure these markers have only recently become available. In a 2026 study, brain-derived p-tau 217 out-performed total p-tau for the classification and staging of AD in a cross-sectional population of 429 Hong Kong Chinese participants^12^. Here, we demonstrated that brain-derived p-tau isoforms could be strong candidates for risk stratification and prognosis of long-term AD risk. We also found that both brain-derived and total p-tau-217/Aβ42 was associated with AD risk, with the ratio between these two proteins being the first blood test for AD diagnosis approved by the Food and Drug Administration in May 2025^30^.

In contrast to previous shorter-term prospective studies with all-cause dementia, not AD, outcomes, we found no association between GFAP or NfL with AD risk ^14,16–18,31^. GFAP is included in the 2024 revised criteria for AD diagnosis and staging, as an inflammatory marker for AD pathogenesis but is non-specific to AD^7^. NfL is a marker of neurodegeneration, with elevated levels typically occurring within 10 years of AD clinical onset^6,14,32^. The lack of association in the current study is consistent with the likelihood that blood was collected in the very early stages of the AD prodrome. The cases and controls were selected from a population-based cohort of ‘healthy’ volunteers with an average age of 63 years at blood collection^20^. This suggests that blood-based biomarkers can predict AD risk in older-middle-aged individuals with healthier lifestyle profiles compared to the general population, and before markers of neurodegeneration are elevated. Reduced Aβ42 is also recognised as a core early biomarker in the 2024 diagnosis and staging criteria. We observed lower relative levels of Aβ42 associated with AD risk, but this was not significant after FDR correction. Grande and colleagues found no association between Aβ42/40 ratio with AD, and hypothesise that this could be due to amyloid concentrations being lower in the blood compared to CSF and the likelihood that mass spectrometry is needed for more accurate detection of Aβ measures^17,33,34^.

Key strengths of this study include the measurement of a range of proteins implicated in CNS processes detectable at low abundance, including brain-derived p-tau isoforms, ascertainment of AD cases over a 25-year follow-up period, and detailed baseline data enabling adjustment for lifestyle, sociodemographic and health-related confounders. By using a panel enriched for established AD markers we provide an independent test of the AUCs in this study and optimism-corrected values in for comparison. This study also has several limitations. The NULISA panel provided relative measurements which limits comparison with studies using absolute quantification. AD cases were identified using hospital and death records which, whilst generally accurate, typically diagnose cases 2-3 years later compared to other sources, such as primary care^35–37^. The AD diagnoses were within a narrow follow-up period, so we were unable to investigate whether the associations varied by different lengths of follow-up. While our sample size is large relative to the length of follow-up, differences in association between the established p-tau markers and novel brain-derived isoforms were small. EPIC-Oxford predominately consists of participants of white ethnic background, and in the current sample only three participants identified as non-white.

In conclusion, our findings suggest that elevation in specific biomarkers, notably brain-derived p-tau 217, precedes clinical detection of AD by at least two decades. This has implications for the design of early interventions and administration of targeted interventions that aim to ameliorate the pathological processes leading to AD.

## Data Availability

The data access policy for the EPIC-Oxford study is available via the study website - https://www.ceu.ox.ac.uk/research/epic-oxford-1/data-access-sharing-and-collaboration

## Acknowledgements

We thank all participants in the EPIC-Oxford cohort for their invaluable contribution. This work uses data provided by patients and collected by the NHS as part of their care and support. We also acknowledge Wolfson Laboratories, Clinical Trial Service Unit and Epidemiological Studies Unit, University of Oxford for the storage and processing of blood samples. For the purpose of open access, the authors have applied a Creative Commons Attribution (CC BY) license to any Author Accepted Manuscript version arising. Thomas Littlejohns and Wenyu Liu had full access to all the data in the study and takes responsibility for the integrity of the data and the accuracy of the data analysis.

## Data sharing

The data access policy for the EPIC-Oxford study is available via the study website (https://www.ceu.ox.ac.uk/research/epic-oxford-1/data-access-sharing-and-collaboration).

