## Supplementary Material for "Blood-based biomarkers and 20-year risk of clinically diagnosed Alzheimer’s disease"

**eTable 1. List of biomarkers on the NULISAseq CNS panel**

| **Biomarker** | **Full protein name** |
| --- | --- |
| Aβ38 | Amyloid-beta precursor protein |
| Aβ40 | Amyloid-beta precursor protein |
| Aβ42 | Amyloid-beta precursor protein |
| ACHE | Acetylcholinesterase |
| AGRN | Agrin |
| ANXA5 | Annexin A5 |
| APOE | Apolipoprotein E |
| APOE4 | Apolipoprotein E isoform 4 |
| ARSA | Arylsulfatase A |
| BACE1 | Beta-secretase 1 |
| BASP1 | Brain acid soluble protein 1 |
| BD p-tau 181 | Brain-derived Tau, phosphorylated at T181 |
| BD p-tau 217 | Brain-derived Tau, phsophorylated at T217 |
| BD p-tau 231 | Brain-derived Tau, phsophorylated at T231 |
| BD Tau | Brain-derived Tau |
| BDNF | Brain-derived neurotrophic factor |
| CALB2 | Calretinin |
| CD40L/TNFSF5 | CD40 ligand |
| CD63 | CD63 antigen |
| CHIT1 | Chitotriosidase-1 |
| CNTN2 | Contactin-2 |
| CRH | Corticoliberin |
| CRP | C-reactive protein |
| CST3 | Cystatin-C |
| CX3CL1/Fractalkine | Fractalkine |
| CXCL1/GROa | Growth-regulated alpha protein |
| DDC | Aromatic-L-Amino-Acid Decarboxylase |
| ENO2 | Gamma-enolase |
| Eotaxin | Eotaxin |
| Eotaxin-3 | C-C motif chemokine 26 |
| FABP3 | Fatty acid-binding protein, heart |
| FCN2 | Ficolin-2 |
| FGF basic | Fibroblast growth factor 2 |
| FOLR1 | Folate receptor alpha |
| GDF15 | Growth/differentiation factor 15 |
| GDI1 | Rab GDP dissociation inhibitor alpha |
| GDNF | Glial cell line-derived neurotrophic factor |
| GFAP | Glial fibrillary acidic protein |
| GM-CSF | Granulocyte-macrophage colony-stimulating factor |
| GOT1 | Aspartate aminotransferase, cytoplasmic |
| HBα1; HBα2 | Hemoglobin subunit alpha 1 \|Hemoglobin subunit alpha 2 |
| HTT | Huntingtin |
| ICAM1 | Intercellular adhesion molecule 1 |
| IFN-gamma | Interferon gamma |
| IGF1R | Insulin-like growth factor 1 Receptor |
| IGFBP7 | Insulin-like growth factor-binding protein 7 |
| IL-1 beta | Interleukin-1 beta |
| IL-10 | Interleukin-10 |
| IL-12p70 | Interleukin 12A \| Interleukin 12B |
| IL-13 | Interleukin-13 |
| IL-15 | Interleukin-15 |
| IL-16 | Pro-interleukin-16 |
| IL-17A | Interleukin-17A |
| IL-18 | Interleukin-18 |
| IL-2 | Interleukin-2 |
| IL-33 | Interleukin-33 |
| IL-4 | Interleukin-4 |
| IL-5 | Interleukin-5 |
| IL-6 | Interleukin-6 |
| IL-6R a | Interleukin-6 receptor subunit alpha |
| IL-7 | Interleukin-7 |
| IL8 | Interleukin-8 |
| IL-9 | Interleukin-9 |
| IP-10 | C-X-C motif chemokine 10 |
| KLK6 | Kallikrein-6 |
| MCP1 | C-C motif chemokine 2 |
| MCP4 | C-C motif chemokine 13 |
| MDC | C-C motif chemokine 22 |
| MDH1 | Malate dehydrogenase, cytoplasmic |
| MIP1a/CCL3 | C-C motif chemokine 3 |
| Mip1b/CCL4 | C-C motif chemokine 4 |
| MME | Neprilysin |
| MSLN | Mesothelin |
| NEFH | Neurofilament heavy polypeptide |
| NfL | Neurofilament light polypeptide |
| NGF | Beta-nerve growth factor |
| NPTX1 | Neuronal pentraxin-1 |
| NPTX2 | Neuronal pentraxin-2 |
| NPTXR | Neuronal pentraxin receptor |
| NPY | Pro-neuropeptide Y |
| NRGN | Neurogranin |
| PARK7 | Parkinson disease protein 7 |
| PDGFRB | Platelet-derived growth factor receptor beta |
| PDLIM5 | PDZ and LIM domain protein 5 |
| PGK1 | Phosphoglycerate kinase 1 |
| PLGF | Placenta growth factor |
| POSTN | Periostin |
| PRDX6 | Peroxiredoxin-6 |
| PSEN1 | Presenilin-1 |
| pSNCA-129 | Phosphorylated alpha-synuclein at serine 129 |
| PTN | Pleiotrophin |
| REST | RE1-silencing transcription factor |
| RUVBL2 | RuvB-like 2 |
| S100A12 | Protein S100-A12 |
| S100B | Protein S100-B |
| SAA1 | Serum amyloid A-1 protein |
| SFRP1 | Secreted frizzled-related protein 1 |
| SFTPD | Pulmonary surfactant-associated protein D |
| SLIT2 | Slit homolog 2 protein |
| SMOC1 | SPARC-related modular calcium-binding protein 1 |
| SNAP25 | Synaptosomal-associated protein 25 |
| SNCAagg | Alpha-synuclein |
| SOD1 | Superoxide dismutase [Cu-Zn] |
| SQSTM1 | Sequestosome-1 |
| sTREM1 | Triggering Receptor Expressed On Myeloid Cells 2 |
| TAFA5 | Chemokine-like protein TAFA-5 |
| TARC/CCL17 | C-C motif chemokine 17 |
| TARDBP | TDP-43 with phosphorylation on serine 409 |
| TDP43 | TAR DNA-binding protein 43 |
| Tie-2/TEK | Angiopoietin-1 receptor |
| TIMP3 | Metalloproteinase inhibitor 3 |
| TNF-a | Tumor necrosis factor |
| Total p-tau 181 | Microtubule-associated protein tau |
| Total p-tau 217 | Microtubule-associated protein tau |
| Total p-tau 231 | Microtubule-associated protein tau |
| Total tau | Microtubule-associated protein tau |
| TREM2 | Triggering receptor expressed on myeloid cells 2 |
| UBB | Polyubiquitin-B |
| UCHL1 | Ubiquitin C-terminal hydrolase L1 |
| VCAM1/CD106 | Vascular cell adhesion protein 1 |
| VEGF R1 | Vascular endothelial growth factor receptor 1 |
| VEGF R2 | Vascular endothelial growth factor receptor 2 |
| VEGF-A | Vascular endothelial growth factor A |
| VEGF-D | Vascular endothelial growth factor D |
| VGF | Neurosecretory protein VGF |
| VILIP-1 | Visinin-like protein 1 |
| YKL40 | Chitinase-3-like protein 1 |
| YWHAG | 14-3-3 protein gamma-B |
| YWHAZ | 14-3-3 protein zeta/delta |
| α-Syn | Alpha-synuclein |
| β-syn | Beta-synuclein |

**eFigure 1. Boxplots demonstrating the distribution of blood-based biomarkers in Alzheimer’s disease cases and matched controls.**

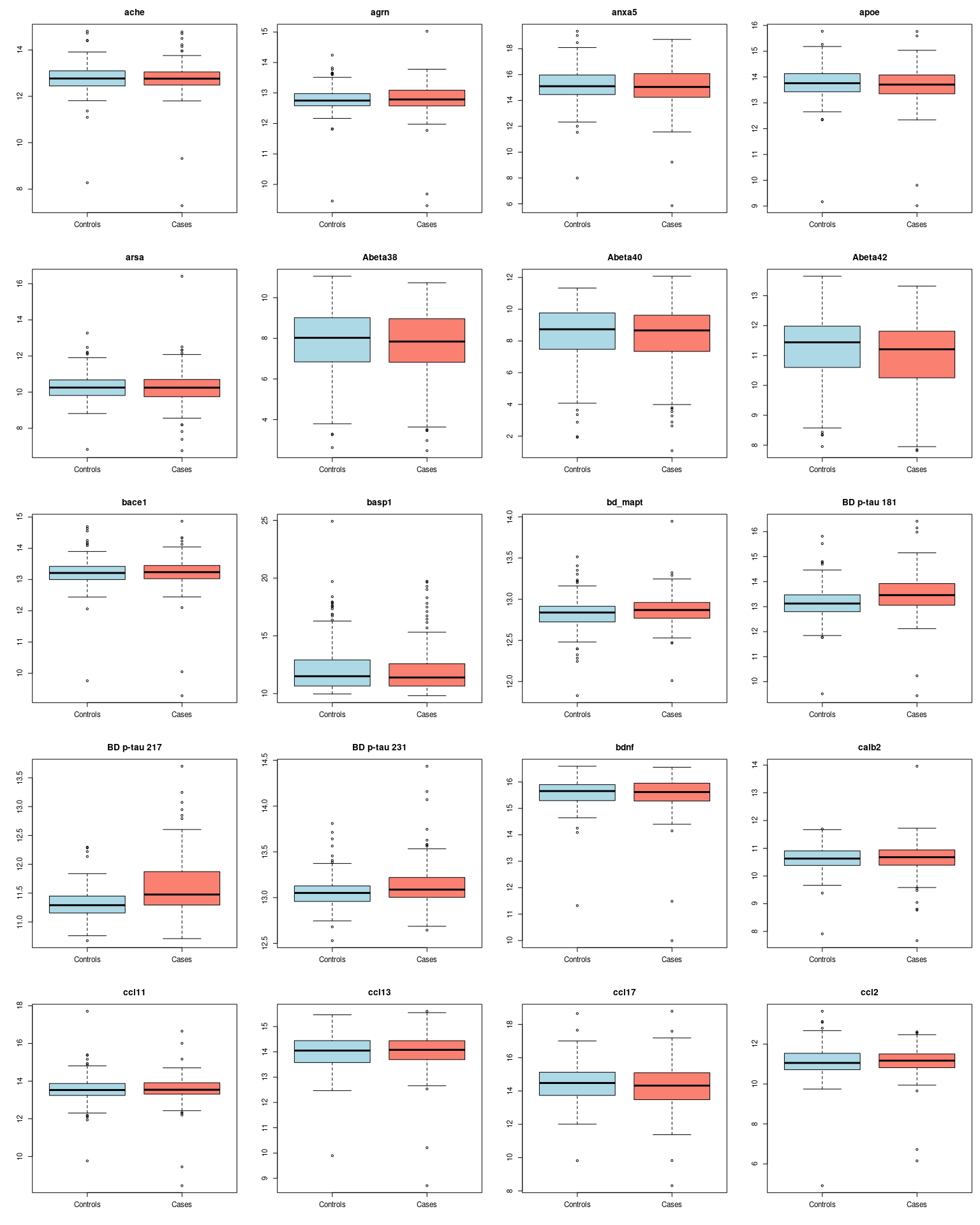

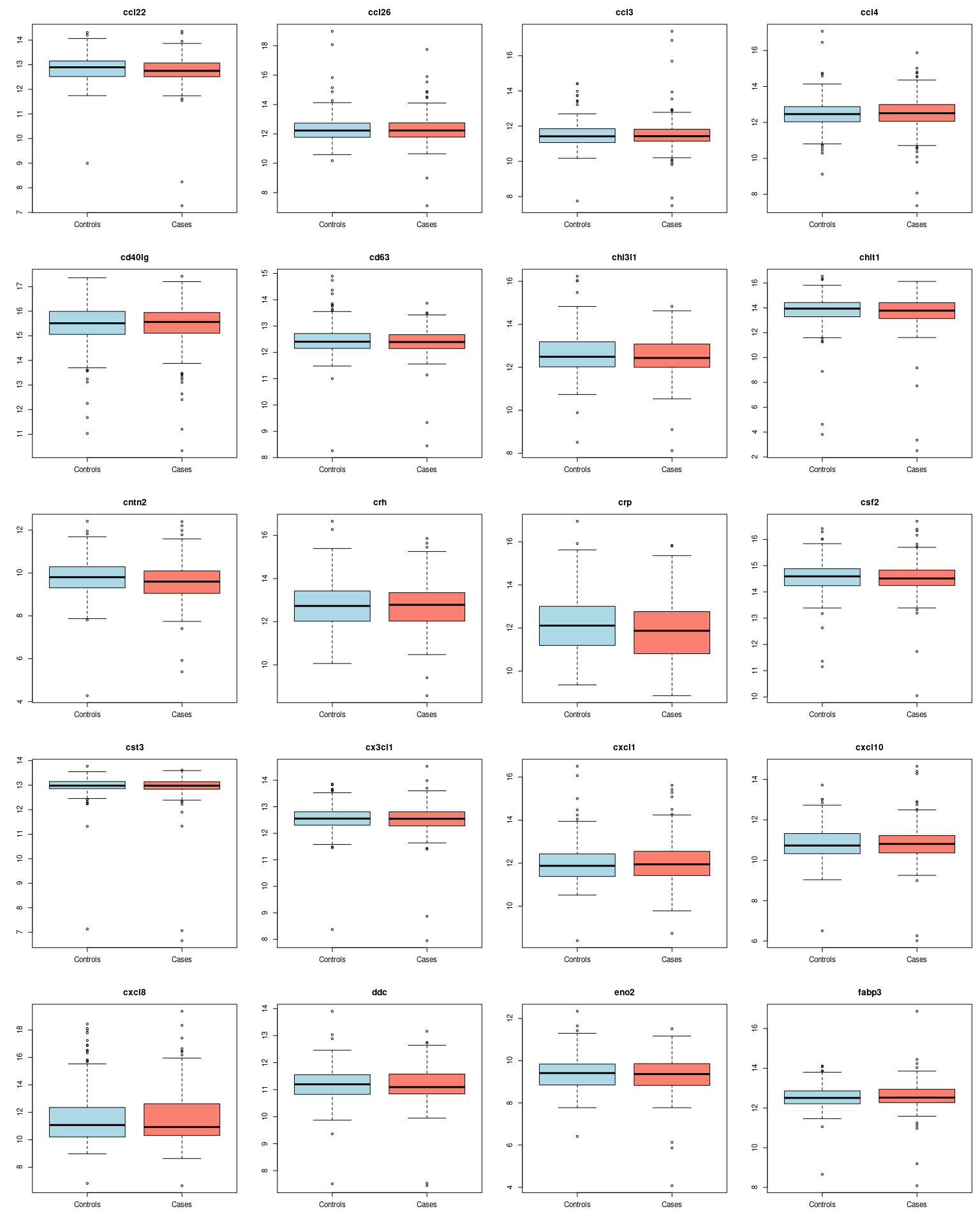

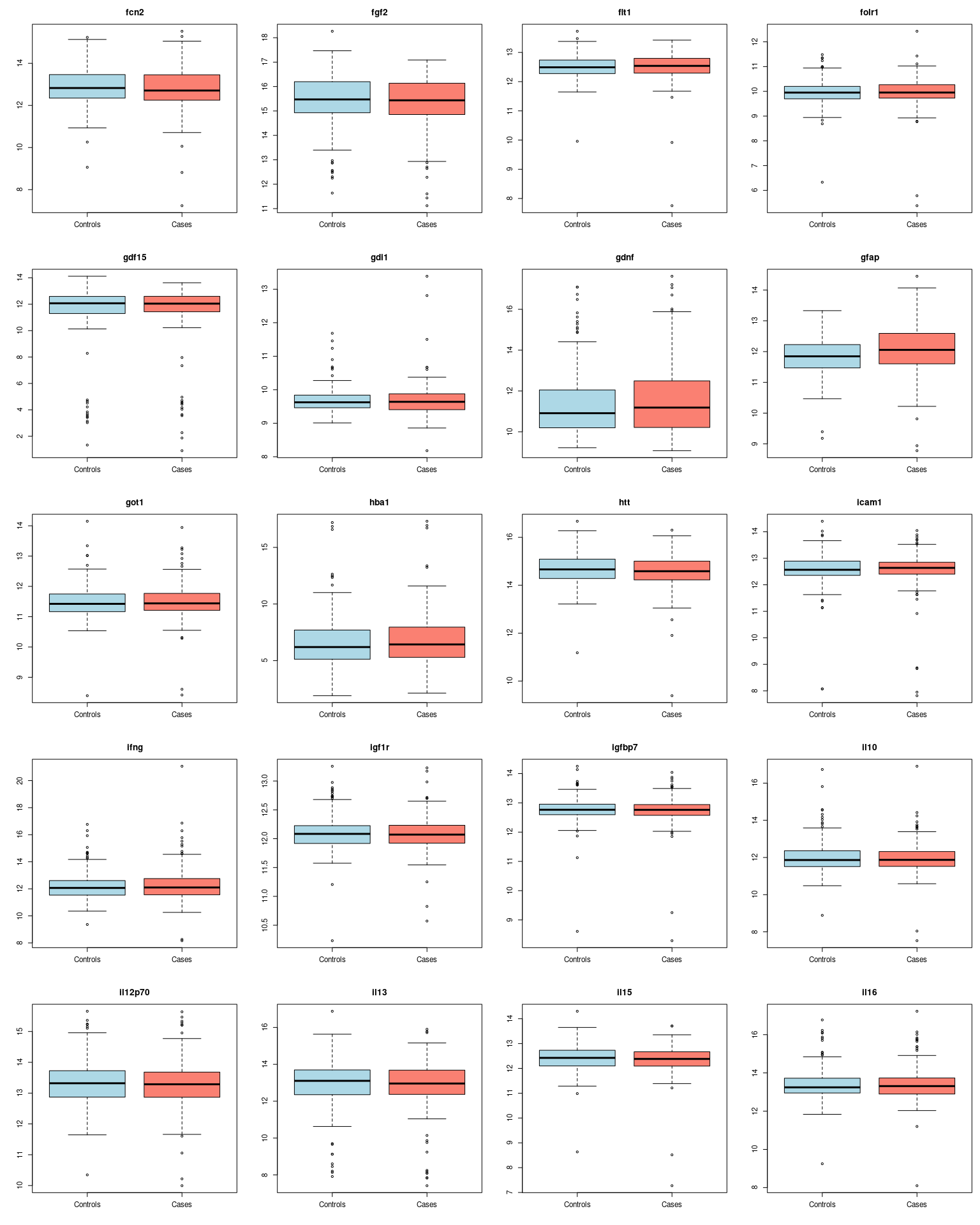

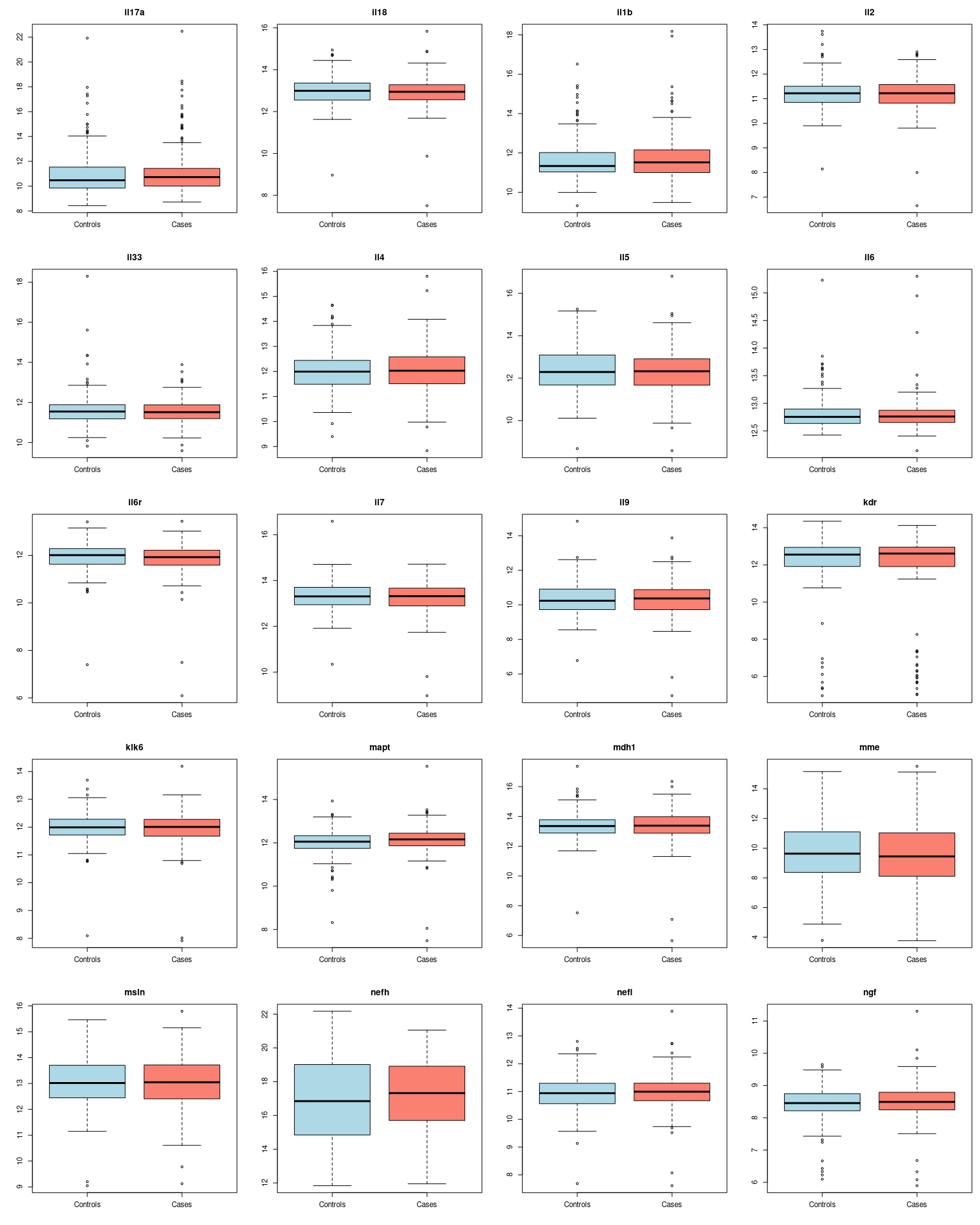

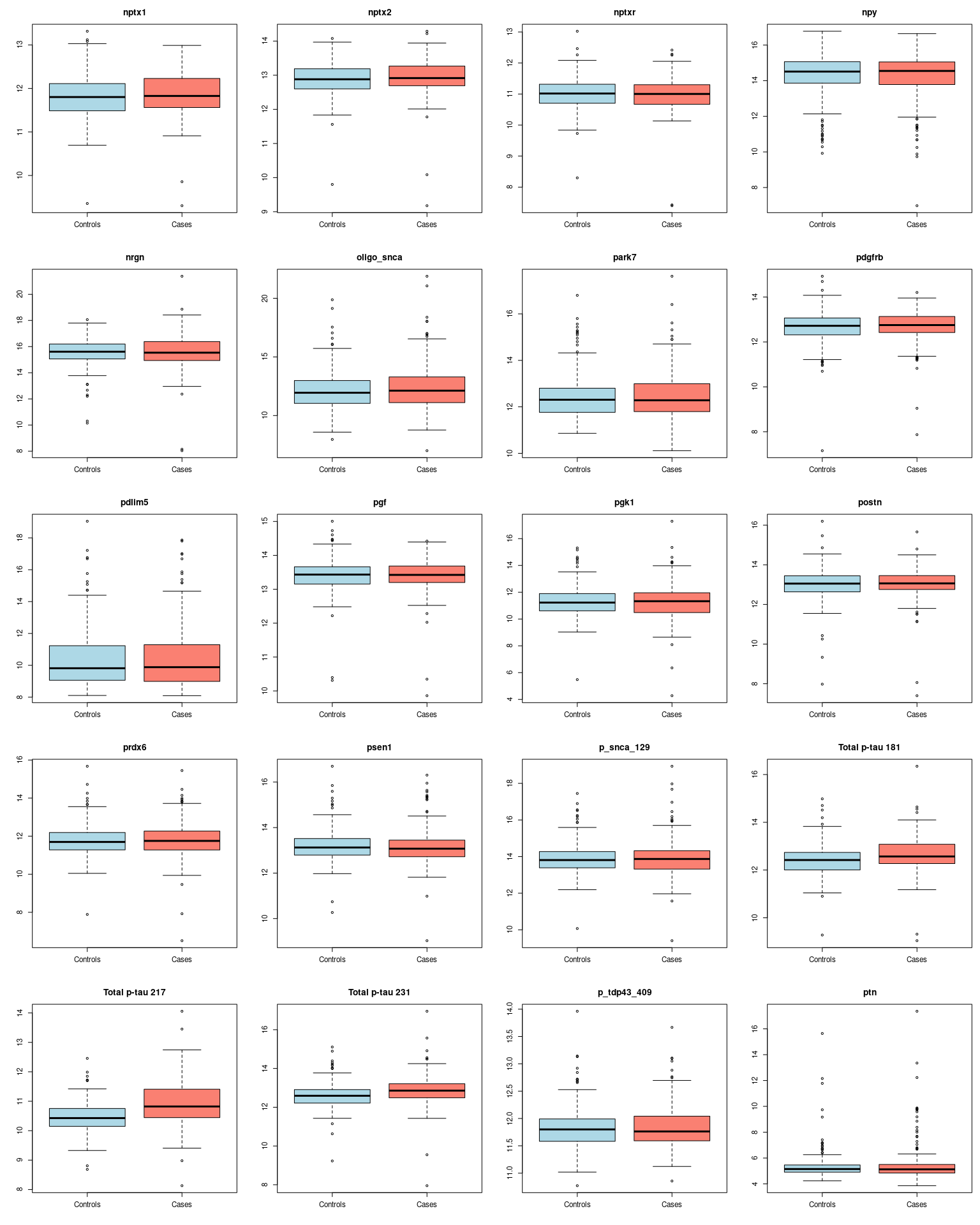

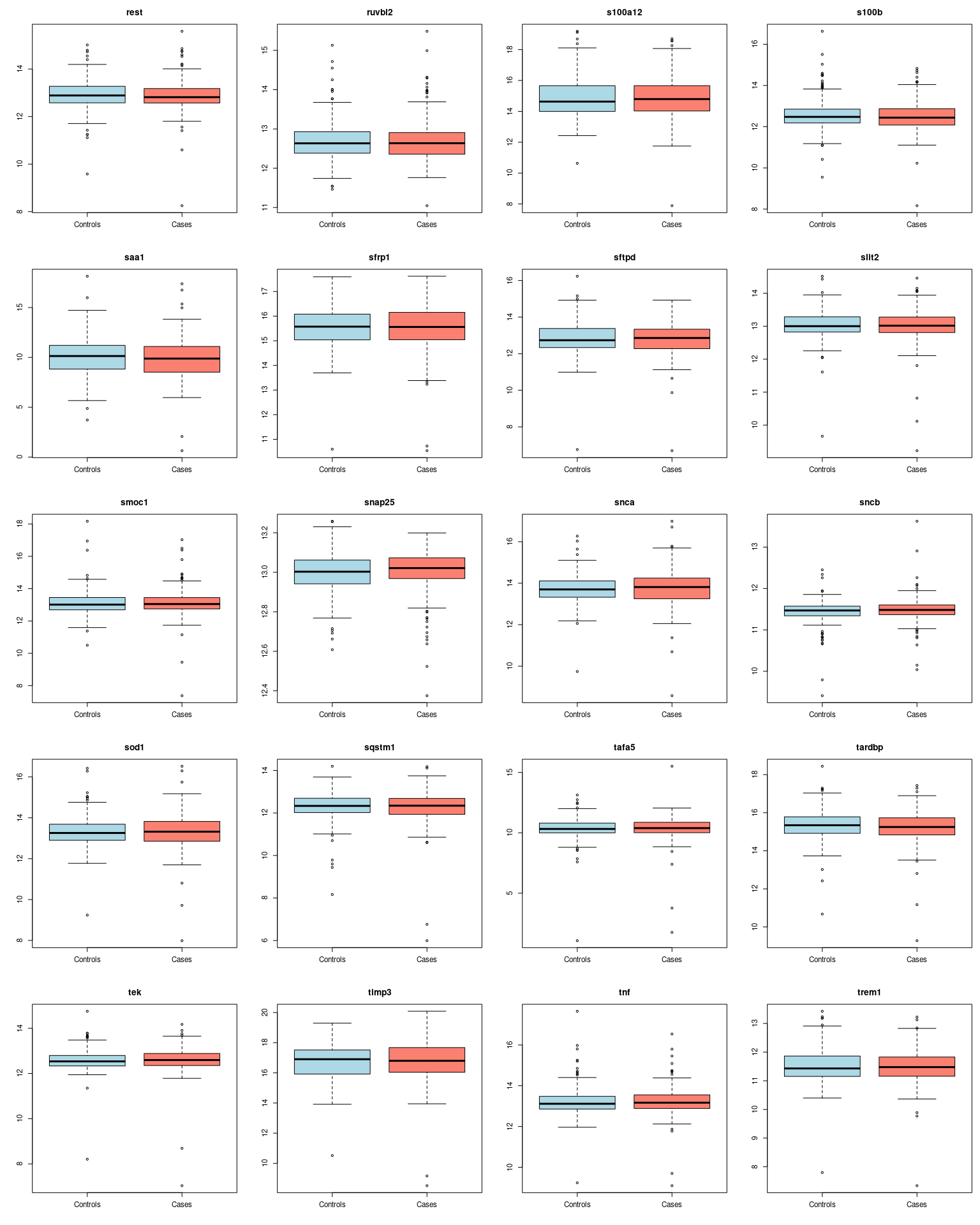

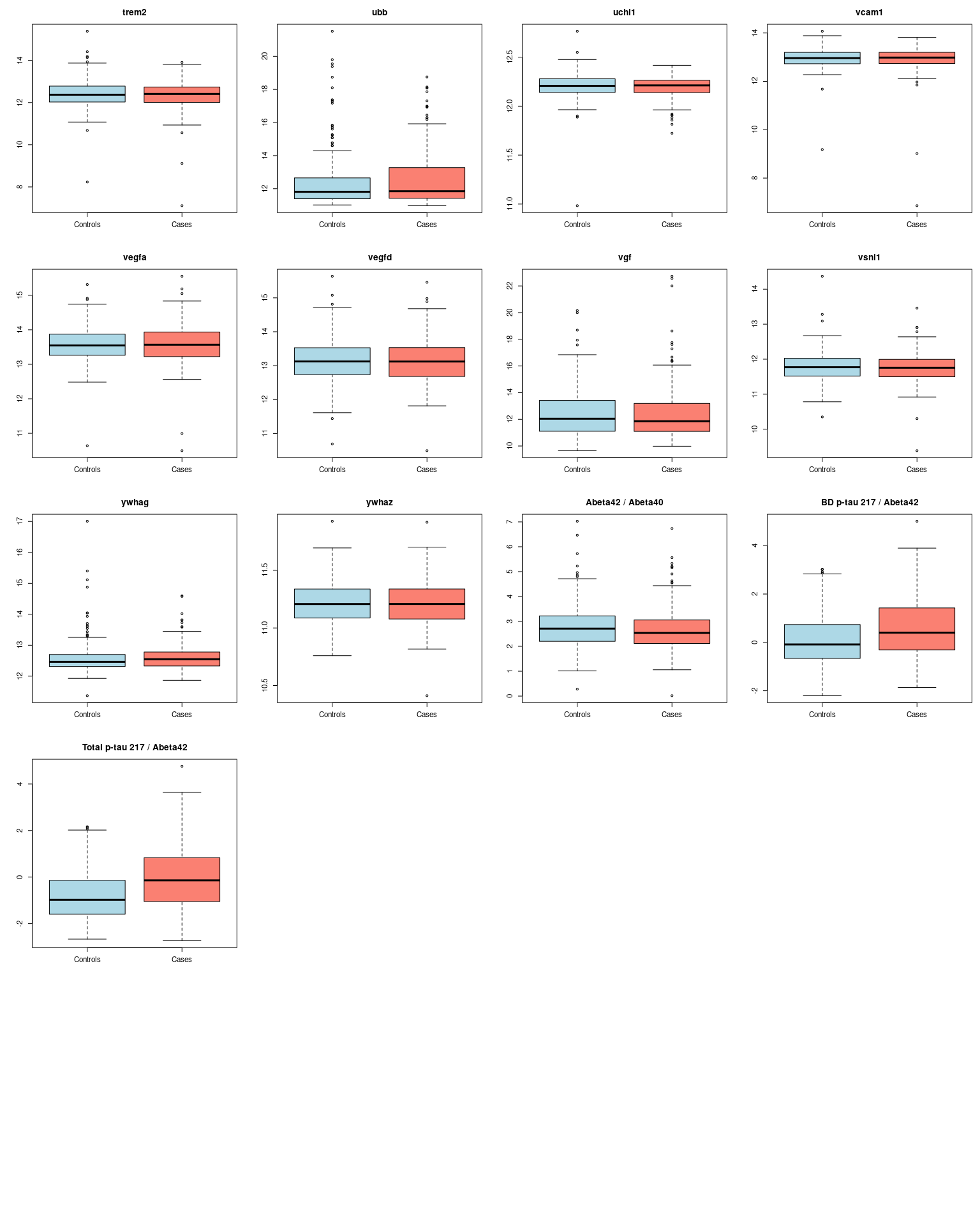

**eFigure 2. Correlation between all 130 blood-based biomarkers on the NULISAseq CNS panel as well as Aβ42/Aβ40, BD p-tau 217 /**
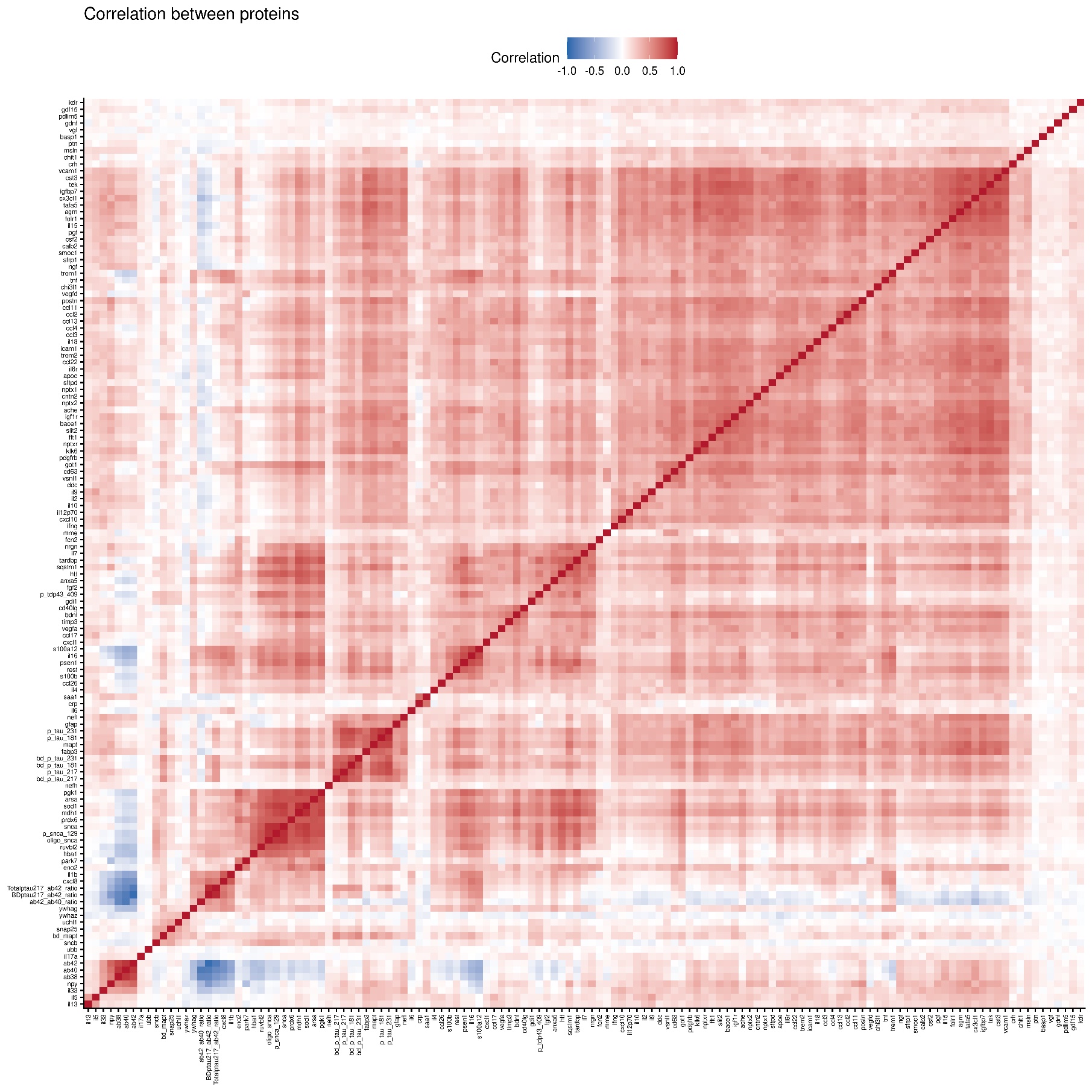
**Aβ42, and total p-tau 217 / Aβ42**

**eTable 2. Odds ratios per 1 standard deviation increase in NPQ (standardised using the control distribution) for association between blood-based biomarkers and Alzheimer’s disease obtained from conditional logistic regression**

|  | **Biomarker only (Model 1)** | | | **Biomarker + APOE4^a^ (Model 2)** | | | **Model 1 + lifestyle^b^** | | | **Model 2 + lifestyle** | | |
| --- | --- | --- | --- | --- | --- | --- | --- | --- | --- | --- | --- | --- |
| **Biomarker** | **OR** | **95% CI** | ***p*** | **OR** | **95% CI** | ***p*** | **OR** | **95% CI** | ***p*** | **OR** | **95%CI** | ***p*** |
| **ACHE** | 0.99 | (0.83, 1.19) | 0.917 | 1.00 | (0.83, 1.21) | 0.999 | 0.96 | (0.79, 1.17) | 0.682 | 0.96 | (0.79, 1.18) | 0.705 |
| **AGRN** | 1.03 | (0.87, 1.22) | 0.732 | 1.02 | (0.85, 1.21) | 0.850 | 1.01 | (0.84, 1.20) | 0.934 | 1.00 | (0.83, 1.20) | 0.960 |
| **ANXA5** | 0.95 | (0.80, 1.13) | 0.573 | 0.97 | (0.81, 1.16) | 0.730 | 0.97 | (0.80, 1.16) | 0.71 | 1.00 | (0.82, 1.21) | 0.985 |
| **APOE** | 0.89 | (0.72, 1.09) | 0.244 | 0.94 | (0.77, 1.16) | 0.591 | 0.88 | (0.71, 1.09) | 0.231 | 0.95 | (0.76, 1.18) | 0.655 |
| **ARSA** | 0.95 | (0.81, 1.13) | 0.590 | 0.99 | (0.83, 1.18) | 0.949 | 0.98 | (0.82, 1.17) | 0.830 | 1.04 | (0.86, 1.25) | 0.707 |
| **AB38** | 0.91 | (0.73, 1.14) | 0.426 | 0.89 | (0.70, 1.12) | 0.323 | 0.92 | (0.72, 1.17) | 0.482 | 0.87 | (0.67, 1.12) | 0.277 |
| **AB40** | 0.93 | (0.75, 1.15) | 0.504 | 0.88 | (0.70, 1.11) | 0.292 | 0.93 | (0.74, 1.17) | 0.521 | 0.87 | (0.68, 1.11) | 0.258 |
| **AB42** | 0.76 | (0.61, 0.94) | 0.012 | 0.73 | (0.58, 0.92) | 0.008 | 0.74 | (0.59, 0.93) | 0.011 | 0.71 | (0.56, 0.91) | 0.007 |
| **BACE1** | 1.01 | (0.84, 1.22) | 0.880 | 0.99 | (0.82, 1.20) | 0.934 | 1.02 | (0.84, 1.25) | 0.831 | 0.97 | (0.79, 1.20) | 0.799 |
| **BASP1** | 0.89 | (0.72, 1.09) | 0.244 | 0.87 | (0.70, 1.07) | 0.189 | 0.89 | (0.72, 1.10) | 0.281 | 0.84 | (0.67, 1.06) | 0.144 |
| **BD MAPT** | 1.40 | (1.12, 1.75) | 0.004 | 1.35 | (1.07, 1.70) | 0.013 | 1.43 | (1.13, 1.82) | 0.003 | 1.36 | (1.06, 1.75) | 0.017 |
| **BD P-TAU 181** | 1.62 | (1.31, 2.01) | <0.001 | 1.49 | (1.20, 1.86) | <0.001 | 1.73 | (1.36, 2.19) | <0.001 | 1.58 | (1.24, 2.01) | <0.001 |
| **BD P-TAU 217** | 2.05 | (1.62, 2.59) | <0.001 | 1.90 | (1.51, 2.40) | <0.001 | 2.29 | (1.75, 3.01) | <0.001 | 2.10 | (1.60, 2.75) | <0.001 |
| **BD P-TAU 231** | 1.44 | (1.17, 1.77) | <0.001 | 1.37 | (1.12, 1.68) | 0.002 | 1.52 | (1.22, 1.90) | <0.001 | 1.44 | (1.15, 1.81) | 0.002 |
| **BDNF** | 0.96 | (0.79, 1.16) | 0.656 | 0.94 | (0.77, 1.14) | 0.523 | 0.97 | (0.80, 1.18) | 0.778 | 0.95 | (0.78, 1.17) | 0.642 |
| **CALB2** | 1.03 | (0.87, 1.22) | 0.743 | 1.04 | (0.87, 1.25) | 0.671 | 1.02 | (0.85, 1.22) | 0.854 | 1.02 | (0.84, 1.24) | 0.812 |
| **CCL11** | 0.99 | (0.81, 1.22) | 0.938 | 1.01 | (0.82, 1.26) | 0.897 | 0.98 | (0.79, 1.22) | 0.878 | 1.01 | (0.80, 1.26) | 0.962 |
| **CCL13** | 1.00 | (0.83, 1.21) | 0.999 | 0.99 | (0.81, 1.21) | 0.938 | 0.99 | (0.81, 1.21) | 0.923 | 0.99 | (0.80, 1.22) | 0.929 |
| **CCL17** | 0.89 | (0.75, 1.06) | 0.178 | 0.92 | (0.77, 1.11) | 0.400 | 0.9 | (0.75, 1.08) | 0.257 | 0.95 | (0.78, 1.15) | 0.572 |
| **CCL2** | 1.00 | (0.82, 1.22) | 0.991 | 0.99 | (0.81, 1.22) | 0.926 | 0.98 | (0.79, 1.21) | 0.816 | 0.97 | (0.78, 1.21) | 0.785 |
| **CCL22** | 0.81 | (0.66, 0.99) | 0.039 | 0.78 | (0.63, 0.98) | 0.030 | 0.81 | (0.65, 1.01) | 0.056 | 0.78 | (0.61, 0.99) | 0.041 |
| **CCL26** | 0.97 | (0.79, 1.20) | 0.802 | 0.94 | (0.76, 1.16) | 0.557 | 0.97 | (0.78, 1.21) | 0.802 | 0.94 | (0.74, 1.19) | 0.598 |
| **CCL3** | 1.03 | (0.88, 1.21) | 0.713 | 1.04 | (0.88, 1.23) | 0.632 | 1.01 | (0.85, 1.19) | 0.942 | 1.01 | (0.85, 1.21) | 0.898 |
| **CCL4** | 1.00 | (0.84, 1.19) | 0.983 | 1.02 | (0.85, 1.23) | 0.829 | 0.97 | (0.80, 1.17) | 0.734 | 0.99 | (0.81, 1.21) | 0.929 |
| **CD40LG** | 0.97 | (0.80, 1.17) | 0.754 | 0.97 | (0.79, 1.18) | 0.749 | 1.00 | (0.82, 1.22) | 0.983 | 1.00 | (0.81, 1.24) | 1.000 |
| **CD63** | 0.86 | (0.70, 1.07) | 0.183 | 0.87 | (0.69, 1.10) | 0.236 | 0.85 | (0.68, 1.08) | 0.189 | 0.84 | (0.65, 1.09) | 0.185 |
| **CHI3L1** | 0.90 | (0.73, 1.10) | 0.301 | 0.93 | (0.75, 1.16) | 0.530 | 0.84 | (0.67, 1.06) | 0.139 | 0.89 | (0.70, 1.13) | 0.349 |
| **CHIT1** | 0.89 | (0.72, 1.08) | 0.240 | 0.88 | (0.71, 1.09) | 0.248 | 0.90 | (0.72, 1.11) | 0.313 | 0.89 | (0.71, 1.11) | 0.296 |
| **CNTN2** | 0.78 | (0.63, 0.96) | 0.019 | 0.78 | (0.62, 0.98) | 0.031 | 0.77 | (0.61, 0.96) | 0.023 | 0.77 | (0.61, 0.98) | 0.033 |
| **CRH** | 1.00 | (0.83, 1.21) | 0.973 | 0.95 | (0.78, 1.16) | 0.646 | 0.99 | (0.81, 1.20) | 0.885 | 0.93 | (0.76, 1.15) | 0.528 |
| **CRP** | 0.84 | (0.69, 1.01) | 0.064 | 0.86 | (0.70, 1.05) | 0.129 | 0.80 | (0.64, 0.99) | 0.043 | 0.85 | (0.67, 1.07) | 0.161 |
| **CSF2** | 0.95 | (0.78, 1.15) | 0.572 | 0.95 | (0.78, 1.15) | 0.584 | 0.93 | (0.76, 1.13) | 0.460 | 0.93 | (0.75, 1.15) | 0.510 |
| **CST3** | 0.94 | (0.79, 1.11) | 0.452 | 0.92 | (0.76, 1.10) | 0.347 | 0.93 | (0.78, 1.11) | 0.418 | 0.91 | (0.76, 1.10) | 0.324 |
| **CX3CL1** | 0.93 | (0.77, 1.12) | 0.437 | 0.92 | (0.76, 1.11) | 0.382 | 0.91 | (0.75, 1.11) | 0.347 | 0.89 | (0.73, 1.10) | 0.288 |
| **CXCL1** | 1.04 | (0.86, 1.25) | 0.700 | 1.05 | (0.86, 1.29) | 0.605 | 1.05 | (0.86, 1.28) | 0.647 | 1.08 | (0.87, 1.34) | 0.478 |
| **CXCL10** | 1.00 | (0.84, 1.20) | 0.991 | 0.99 | (0.82, 1.20) | 0.899 | 0.96 | (0.79, 1.16) | 0.651 | 0.93 | (0.76, 1.15) | 0.508 |
| **CXCL8** | 1.06 | (0.86, 1.32) | 0.560 | 1.07 | (0.85, 1.34) | 0.570 | 1.08 | (0.86, 1.36) | 0.494 | 1.08 | (0.85, 1.37) | 0.533 |
| **DDC** | 0.97 | (0.80, 1.17) | 0.722 | 0.97 | (0.79, 1.18) | 0.756 | 0.95 | (0.78, 1.17) | 0.634 | 0.95 | (0.77, 1.17) | 0.615 |
| **ENO2** | 0.94 | (0.76, 1.16) | 0.544 | 0.93 | (0.75, 1.17) | 0.541 | 0.94 | (0.75, 1.19) | 0.607 | 0.94 | (0.73, 1.20) | 0.603 |
| **FABP3** | 1.06 | (0.89, 1.26) | 0.511 | 1.07 | (0.89, 1.28) | 0.468 | 1.02 | (0.85, 1.22) | 0.862 | 1.04 | (0.86, 1.25) | 0.718 |
| **FCN2** | 0.82 | (0.61, 1.09) | 0.162 | 0.80 | (0.59, 1.08) | 0.140 | 0.79 | (0.59, 1.07) | 0.135 | 0.77 | (0.56, 1.07) | 0.119 |
| **FGF2** | 0.95 | (0.79, 1.15) | 0.622 | 0.96 | (0.78, 1.17) | 0.681 | 0.94 | (0.77, 1.16) | 0.560 | 0.94 | (0.75, 1.17) | 0.583 |
| **FLT1** | 0.96 | (0.81, 1.14) | 0.621 | 0.94 | (0.79, 1.13) | 0.520 | 0.95 | (0.80, 1.14) | 0.602 | 0.93 | (0.77, 1.13) | 0.472 |
| **FOLR1** | 1.03 | (0.86, 1.22) | 0.782 | 1.05 | (0.87, 1.26) | 0.623 | 1.01 | (0.83, 1.22) | 0.936 | 1.03 | (0.85, 1.26) | 0.764 |
| **GDF15** | 1.00 | (0.82, 1.22) | 0.991 | 0.98 | (0.80, 1.21) | 0.872 | 0.96 | (0.78, 1.18) | 0.702 | 0.94 | (0.75, 1.18) | 0.590 |
| **GDI1** | 1.01 | (0.85, 1.20) | 0.873 | 1.02 | (0.86, 1.23) | 0.790 | 1.04 | (0.87, 1.24) | 0.699 | 1.05 | (0.87, 1.27) | 0.588 |
| **GDNF** | 1.21 | (1.01, 1.44) | 0.036 | 1.20 | (0.99, 1.44) | 0.062 | 1.18 | (0.98, 1.43) | 0.079 | 1.17 | (0.96, 1.42) | 0.126 |
| **GFAP** | 1.39 | (1.15, 1.70) | 0.001 | 1.29 | (1.05, 1.58) | 0.013 | 1.44 | (1.16, 1.79) | 0.001 | 1.30 | (1.03, 1.63) | 0.024 |
| **GOT1** | 1.03 | (0.85, 1.25) | 0.750 | 1.01 | (0.83, 1.23) | 0.899 | 1.05 | (0.86, 1.28) | 0.656 | 1.03 | (0.84, 1.27) | 0.780 |
| **HBA1** | 1.15 | (0.95, 1.41) | 0.156 | 1.21 | (0.98, 1.50) | 0.078 | 1.18 | (0.96, 1.46) | 0.122 | 1.26 | (1.00, 1.58) | 0.048 |
| **HTT** | 0.92 | (0.77, 1.11) | 0.377 | 0.93 | (0.77, 1.13) | 0.475 | 0.94 | (0.78, 1.14) | 0.547 | 0.97 | (0.79, 1.18) | 0.749 |
| **ICAM1** | 0.96 | (0.80, 1.16) | 0.688 | 0.95 | (0.78, 1.15) | 0.604 | 0.96 | (0.79, 1.16) | 0.672 | 0.96 | (0.79, 1.17) | 0.683 |
| **IFNG** | 1.09 | (0.91, 1.31) | 0.328 | 1.10 | (0.90, 1.33) | 0.354 | 1.07 | (0.88, 1.29) | 0.522 | 1.08 | (0.87, 1.33) | 0.497 |
| **IGF1R** | 0.94 | (0.76, 1.17) | 0.585 | 0.94 | (0.74, 1.18) | 0.582 | 0.93 | (0.74, 1.17) | 0.535 | 0.91 | (0.71, 1.17) | 0.479 |
| **IGFBP7** | 0.99 | (0.83, 1.18) | 0.914 | 0.97 | (0.81, 1.17) | 0.777 | 0.98 | (0.81, 1.18) | 0.812 | 0.95 | (0.78, 1.16) | 0.620 |
| **IL10** | 0.92 | (0.76, 1.12) | 0.413 | 0.91 | (0.74, 1.12) | 0.357 | 0.89 | (0.72, 1.09) | 0.263 | 0.87 | (0.70, 1.09) | 0.221 |
| **IL12P70** | 0.98 | (0.81, 1.18) | 0.825 | 1.00 | (0.82, 1.23) | 0.972 | 0.97 | (0.79, 1.19) | 0.784 | 0.99 | (0.79, 1.22) | 0.892 |
| **IL13** | 0.97 | (0.81, 1.17) | 0.776 | 0.94 | (0.77, 1.14) | 0.540 | 0.95 | (0.78, 1.15) | 0.598 | 0.91 | (0.74, 1.12) | 0.375 |
| **IL15** | 0.93 | (0.77, 1.11) | 0.407 | 0.93 | (0.77, 1.12) | 0.432 | 0.90 | (0.74, 1.10) | 0.307 | 0.90 | (0.73, 1.10) | 0.303 |
| **IL16** | 1.01 | (0.84, 1.22) | 0.906 | 1.02 | (0.84, 1.24) | 0.862 | 1.04 | (0.85, 1.27) | 0.714 | 1.06 | (0.86, 1.31) | 0.586 |
| **IL17A** | 1.10 | (0.91, 1.32) | 0.342 | 1.14 | (0.93, 1.40) | 0.201 | 1.12 | (0.92, 1.36) | 0.273 | 1.16 | (0.94, 1.43) | 0.169 |
| **IL18** | 0.92 | (0.75, 1.12) | 0.402 | 0.96 | (0.78, 1.17) | 0.667 | 0.93 | (0.75, 1.15) | 0.480 | 0.97 | (0.78, 1.22) | 0.823 |
| **IL1B** | 1.11 | (0.92, 1.35) | 0.267 | 1.16 | (0.95, 1.43) | 0.144 | 1.17 | (0.95, 1.44) | 0.136 | 1.23 | (0.99, 1.54) | 0.067 |
| **IL2** | 0.95 | (0.78, 1.14) | 0.559 | 0.94 | (0.77, 1.15) | 0.546 | 0.94 | (0.77, 1.15) | 0.540 | 0.94 | (0.76, 1.16) | 0.552 |
| **IL33** | 0.85 | (0.67, 1.08) | 0.187 | 0.84 | (0.65, 1.09) | 0.196 | 0.80 | (0.62, 1.03) | 0.085 | 0.79 | (0.59, 1.06) | 0.114 |
| **IL4** | 1.05 | (0.87, 1.27) | 0.618 | 1.01 | (0.83, 1.24) | 0.898 | 1.07 | (0.87, 1.31) | 0.545 | 1.03 | (0.83, 1.29) | 0.766 |
| **IL5** | 0.95 | (0.79, 1.14) | 0.567 | 0.97 | (0.80, 1.17) | 0.759 | 0.95 | (0.78, 1.14) | 0.569 | 0.98 | (0.80, 1.20) | 0.857 |
| **IL6** | 0.97 | (0.80, 1.16) | 0.718 | 0.93 | (0.76, 1.13) | 0.442 | 0.99 | (0.81, 1.21) | 0.915 | 0.97 | (0.78, 1.19) | 0.752 |
| **IL6R** | 0.93 | (0.78, 1.11) | 0.431 | 0.90 | (0.74, 1.09) | 0.266 | 0.90 | (0.74, 1.08) | 0.264 | 0.87 | (0.71, 1.06) | 0.166 |
| **IL7** | 0.91 | (0.76, 1.09) | 0.311 | 0.91 | (0.75, 1.10) | 0.330 | 0.93 | (0.77, 1.12) | 0.461 | 0.93 | (0.76, 1.13) | 0.461 |
| **IL9** | 0.97 | (0.80, 1.17) | 0.726 | 0.99 | (0.81, 1.20) | 0.909 | 0.95 | (0.78, 1.16) | 0.590 | 0.96 | (0.78, 1.19) | 0.723 |
| **KDR** | 0.88 | (0.74, 1.04) | 0.123 | 0.91 | (0.76, 1.08) | 0.285 | 0.87 | (0.72, 1.05) | 0.136 | 0.91 | (0.74, 1.10) | 0.321 |
| **KLK6** | 0.97 | (0.82, 1.15) | 0.747 | 0.96 | (0.80, 1.15) | 0.661 | 0.95 | (0.79, 1.15) | 0.615 | 0.93 | (0.76, 1.12) | 0.434 |
| **MAPT** | 1.29 | (1.04, 1.58) | 0.018 | 1.23 | (1.00, 1.52) | 0.054 | 1.26 | (1.01, 1.57) | 0.039 | 1.20 | (0.96, 1.50) | 0.110 |
| **MDH1** | 1.01 | (0.85, 1.19) | 0.931 | 1.04 | (0.87, 1.24) | 0.666 | 1.03 | (0.86, 1.23) | 0.727 | 1.08 | (0.90, 1.31) | 0.410 |
| **MME** | 0.93 | (0.77, 1.12) | 0.444 | 0.91 | (0.74, 1.12) | 0.362 | 0.92 | (0.75, 1.13) | 0.445 | 0.91 | (0.73, 1.14) | 0.421 |
| **MSLN** | 1.02 | (0.84, 1.24) | 0.820 | 1.08 | (0.88, 1.32) | 0.477 | 1.03 | (0.84, 1.26) | 0.777 | 1.09 | (0.88, 1.35) | 0.430 |
| **NEFH** | 1.17 | (0.94, 1.45) | 0.172 | 1.17 | (0.93, 1.48) | 0.181 | 1.19 | (0.94, 1.50) | 0.156 | 1.20 | (0.93, 1.54) | 0.153 |
| **NEFL** | 1.05 | (0.86, 1.28) | 0.612 | 1.09 | (0.88, 1.34) | 0.439 | 1.04 | (0.84, 1.30) | 0.700 | 1.07 | (0.85, 1.35) | 0.556 |
| **NGF** | 1.11 | (0.91, 1.35) | 0.323 | 1.09 | (0.88, 1.34) | 0.429 | 1.09 | (0.88, 1.35) | 0.420 | 1.07 | (0.86, 1.34) | 0.533 |
| **NPTX1** | 1.10 | (0.90, 1.35) | 0.332 | 1.04 | (0.84, 1.28) | 0.725 | 1.10 | (0.89, 1.37) | 0.366 | 1.01 | (0.80, 1.27) | 0.932 |
| **NPTX2** | 1.16 | (0.95, 1.43) | 0.153 | 1.09 | (0.88, 1.35) | 0.410 | 1.16 | (0.93, 1.44) | 0.186 | 1.07 | (0.86, 1.34) | 0.548 |
| **NPTXR** | 0.93 | (0.78, 1.12) | 0.459 | 0.92 | (0.76, 1.11) | 0.371 | 0.90 | (0.73, 1.09) | 0.276 | 0.85 | (0.69, 1.05) | 0.137 |
| **NPY** | 0.94 | (0.76, 1.18) | 0.606 | 0.91 | (0.72, 1.14) | 0.401 | 0.92 | (0.73, 1.17) | 0.498 | 0.88 | (0.68, 1.12) | 0.294 |
| **NRGN** | 1.01 | (0.85, 1.21) | 0.884 | 0.99 | (0.82, 1.19) | 0.925 | 1.04 | (0.86, 1.25) | 0.720 | 1.01 | (0.83, 1.24) | 0.895 |
| **OLIGO_SNCA** | 1.11 | (0.93, 1.32) | 0.254 | 1.14 | (0.95, 1.37) | 0.168 | 1.15 | (0.95, 1.40) | 0.158 | 1.22 | (0.99, 1.50) | 0.062 |
| **PARK7** | 1.08 | (0.83, 1.41) | 0.560 | 1.11 | (0.84, 1.46) | 0.458 | 1.10 | (0.83, 1.45) | 0.518 | 1.16 | (0.86, 1.55) | 0.325 |
| **PDGFRB** | 0.99 | (0.81, 1.22) | 0.960 | 1.01 | (0.82, 1.25) | 0.919 | 0.95 | (0.77, 1.18) | 0.663 | 0.96 | (0.77, 1.21) | 0.748 |
| **PDLIM5** | 1.03 | (0.85, 1.24) | 0.777 | 1.06 | (0.87, 1.30) | 0.567 | 1.05 | (0.86, 1.29) | 0.616 | 1.09 | (0.88, 1.36) | 0.429 |
| **PGF** | 0.99 | (0.81, 1.22) | 0.927 | 0.94 | (0.76, 1.17) | 0.583 | 0.99 | (0.80, 1.23) | 0.908 | 0.93 | (0.74, 1.17) | 0.558 |
| **PGK1** | 1.00 | (0.83, 1.20) | 0.981 | 1.02 | (0.84, 1.23) | 0.872 | 1.03 | (0.85, 1.25) | 0.765 | 1.06 | (0.87, 1.30) | 0.555 |
| **POSTN** | 1.02 | (0.83, 1.24) | 0.881 | 0.98 | (0.80, 1.21) | 0.881 | 0.99 | (0.80, 1.23) | 0.949 | 0.95 | (0.76, 1.19) | 0.673 |
| **PRDX6** | 0.99 | (0.82, 1.20) | 0.946 | 1.05 | (0.86, 1.27) | 0.647 | 1.01 | (0.82, 1.23) | 0.951 | 1.10 | (0.88, 1.36) | 0.405 |
| **PSEN1** | 0.98 | (0.82, 1.18) | 0.828 | 0.98 | (0.81, 1.19) | 0.854 | 1.01 | (0.83, 1.23) | 0.910 | 1.03 | (0.83, 1.26) | 0.803 |
| **P-SNCA 129** | 1.03 | (0.86, 1.23) | 0.753 | 1.07 | (0.89, 1.28) | 0.482 | 1.05 | (0.87, 1.27) | 0.596 | 1.12 | (0.92, 1.37) | 0.264 |
| **Total P-TAU 181** | 1.41 | (1.14, 1.74) | 0.001 | 1.34 | (1.08, 1.66) | 0.007 | 1.4 | (1.12, 1.75) | 0.004 | 1.33 | (1.05, 1.67) | 0.016 |
| **Total P-TAU 217** | 1.94 | (1.56, 2.41) | <0.001 | 1.78 | (1.43, 2.22) | <0.001 | 2.08 | (1.63, 2.67) | <0.001 | 1.89 | (1.47, 2.42) | <0.001 |
| **Total P-TAU 231** | 1.45 | (1.18, 1.78) | <0.001 | 1.38 | (1.12, 1.70) | 0.003 | 1.48 | (1.18, 1.85) | 0.001 | 1.40 | (1.11, 1.76) | 0.004 |
| **P-TDP43 409** | 1.02 | (0.84, 1.25) | 0.835 | 1.04 | (0.84, 1.28) | 0.745 | 1.08 | (0.86, 1.36) | 0.505 | 1.14 | (0.89, 1.45) | 0.316 |
| **PTN** | 1.10 | (0.92, 1.32) | 0.275 | 1.07 | (0.89, 1.28) | 0.492 | 1.10 | (0.92, 1.32) | 0.292 | 1.06 | (0.88, 1.28) | 0.533 |
| **REST** | 0.97 | (0.80, 1.17) | 0.723 | 0.97 | (0.80, 1.17) | 0.729 | 1.00 | (0.82, 1.22) | 0.974 | 1.02 | (0.82, 1.26) | 0.887 |
| **RUVBL2** | 1.04 | (0.86, 1.25) | 0.702 | 1.07 | (0.88, 1.31) | 0.478 | 1.09 | (0.88, 1.34) | 0.416 | 1.18 | (0.94, 1.47) | 0.155 |
| **S100A12** | 1.07 | (0.89, 1.30) | 0.476 | 1.09 | (0.89, 1.32) | 0.420 | 1.11 | (0.91, 1.37) | 0.309 | 1.13 | (0.92, 1.40) | 0.252 |
| **S100B** | 0.89 | (0.74, 1.08) | 0.236 | 0.88 | (0.71, 1.07) | 0.198 | 0.89 | (0.73, 1.09) | 0.253 | 0.88 | (0.71, 1.09) | 0.233 |
| **SAA1** | 0.91 | (0.75, 1.10) | 0.328 | 0.89 | (0.73, 1.08) | 0.226 | 0.88 | (0.71, 1.09) | 0.238 | 0.86 | (0.69, 1.08) | 0.200 |
| **SFRP1** | 0.98 | (0.82, 1.17) | 0.799 | 0.93 | (0.77, 1.12) | 0.452 | 0.99 | (0.83, 1.19) | 0.947 | 0.94 | (0.77, 1.14) | 0.534 |
| **SFTPD** | 0.95 | (0.77, 1.17) | 0.604 | 0.91 | (0.73, 1.14) | 0.421 | 0.96 | (0.76, 1.20) | 0.69 | 0.91 | (0.72, 1.16) | 0.457 |
| **SLIT2** | 0.94 | (0.79, 1.13) | 0.524 | 0.91 | (0.75, 1.10) | 0.309 | 0.96 | (0.79, 1.15) | 0.632 | 0.91 | (0.75, 1.11) | 0.362 |
| **SMOC1** | 1.04 | (0.86, 1.26) | 0.709 | 1.01 | (0.82, 1.24) | 0.943 | 1.03 | (0.85, 1.26) | 0.751 | 1.00 | (0.80, 1.23) | 0.964 |
| **SNAP25** | 1.06 | (0.88, 1.28) | 0.530 | 0.96 | (0.78, 1.17) | 0.670 | 1.09 | (0.90, 1.33) | 0.365 | 0.98 | (0.79, 1.20) | 0.823 |
| **SNCA** | 1.06 | (0.89, 1.26) | 0.513 | 1.08 | (0.91, 1.30) | 0.383 | 1.08 | (0.89, 1.30) | 0.428 | 1.13 | (0.93, 1.37) | 0.233 |
| **SNCB** | 1.15 | (0.95, 1.38) | 0.156 | 1.14 | (0.94, 1.39) | 0.196 | 1.17 | (0.96, 1.43) | 0.130 | 1.18 | (0.95, 1.46) | 0.138 |
| **SOD1** | 1.04 | (0.87, 1.23) | 0.679 | 1.07 | (0.90, 1.28) | 0.452 | 1.05 | (0.88, 1.26) | 0.596 | 1.1 | (0.91, 1.33) | 0.319 |
| **SQSTM1** | 0.98 | (0.81, 1.17) | 0.806 | 0.97 | (0.80, 1.18) | 0.772 | 0.98 | (0.81, 1.20) | 0.879 | 0.99 | (0.81, 1.22) | 0.947 |
| **TAFA5** | 1.03 | (0.86, 1.25) | 0.727 | 1.03 | (0.85, 1.25) | 0.756 | 1.02 | (0.84, 1.25) | 0.832 | 1.01 | (0.83, 1.24) | 0.894 |
| **TARDBP** | 0.93 | (0.77, 1.12) | 0.449 | 0.94 | (0.77, 1.14) | 0.523 | 0.95 | (0.78, 1.16) | 0.638 | 0.97 | (0.79, 1.20) | 0.811 |
| **TEK** | 0.99 | (0.83, 1.18) | 0.897 | 0.97 | (0.81, 1.17) | 0.764 | 0.98 | (0.81, 1.18) | 0.830 | 0.96 | (0.79, 1.17) | 0.672 |
| **TIMP3** | 1.05 | (0.87, 1.27) | 0.624 | 1.02 | (0.84, 1.24) | 0.844 | 1.06 | (0.86, 1.30) | 0.608 | 1.03 | (0.83, 1.28) | 0.808 |
| **TNF** | 1.01 | (0.83, 1.22) | 0.945 | 1.03 | (0.84, 1.26) | 0.792 | 0.99 | (0.81, 1.22) | 0.948 | 1.02 | (0.82, 1.27) | 0.864 |
| **TREM1** | 0.93 | (0.76, 1.15) | 0.503 | 0.95 | (0.76, 1.18) | 0.628 | 0.91 | (0.73, 1.15) | 0.431 | 0.94 | (0.75, 1.20) | 0.634 |
| **TREM2** | 0.95 | (0.78, 1.16) | 0.626 | 0.96 | (0.78, 1.18) | 0.674 | 0.97 | (0.79, 1.19) | 0.766 | 0.98 | (0.78, 1.22) | 0.824 |
| **UBB** | 1.07 | (0.88, 1.29) | 0.484 | 1.07 | (0.87, 1.31) | 0.507 | 1.07 | (0.87, 1.30) | 0.532 | 1.05 | (0.84, 1.30) | 0.690 |
| **UCHL1** | 0.95 | (0.76, 1.18) | 0.634 | 0.97 | (0.77, 1.23) | 0.805 | 0.95 | (0.76, 1.19) | 0.634 | 0.96 | (0.76, 1.22) | 0.766 |
| **VCAM1** | 0.96 | (0.82, 1.13) | 0.615 | 0.94 | (0.79, 1.11) | 0.441 | 0.93 | (0.79, 1.11) | 0.439 | 0.90 | (0.74, 1.08) | 0.262 |
| **VEGFA** | 0.98 | (0.81, 1.19) | 0.853 | 0.96 | (0.78, 1.17) | 0.683 | 1.00 | (0.81, 1.22) | 0.962 | 0.97 | (0.78, 1.20) | 0.767 |
| **VEGFD** | 0.94 | (0.77, 1.14) | 0.533 | 0.96 | (0.78, 1.18) | 0.691 | 0.90 | (0.73, 1.12) | 0.351 | 0.92 | (0.74, 1.15) | 0.48 |
| **VGF** | 0.98 | (0.82, 1.18) | 0.870 | 0.97 | (0.80, 1.19) | 0.789 | 1.01 | (0.83, 1.23) | 0.934 | 1.00 | (0.80, 1.23) | 0.969 |
| **VSNL1** | 0.94 | (0.77, 1.16) | 0.564 | 0.93 | (0.75, 1.15) | 0.508 | 0.95 | (0.76, 1.19) | 0.657 | 0.93 | (0.74, 1.17) | 0.536 |
| **YWHAG** | 1.02 | (0.82, 1.27) | 0.857 | 1.05 | (0.83, 1.33) | 0.668 | 1.01 | (0.81, 1.27) | 0.911 | 1.04 | (0.81, 1.32) | 0.764 |
| **YWHAZ** | 0.98 | (0.80, 1.19) | 0.826 | 1.00 | (0.82, 1.23) | 0.973 | 1.02 | (0.83, 1.25) | 0.855 | 1.06 | (0.85, 1.31) | 0.628 |
| **Aβ42/Aβ40** | 0.85 | (0.69, 1.05) | 0.129 | 0.90 | (0.72, 1.12) | 0.349 | 0.83 | (0.66, 1.04) | 0.098 | 0.89 | (0.70, 1.13) | 0.325 |
| **BD p-tau 217 /Aβ42** | 1.75 | (1.39, 2.21) | <0.001 | 1.74 | (1.37, 2.22) | <0.001 | 1.87 | (1.45, 2.41) | <0.001 | 1.84 | (1.42, 2.39) | <0.001 |
| **Total p-tau 217 /Aβ42** | 1.91 | (1.52, 2.41) | <0.001 | 1.87 | (1.47, 2.37) | <0.001 | 2.08 | (1.61, 2.69) | <0.001 | 2.01 | (1.54, 2.61) | <0.001 |

Abbreviations: CI, Confidence Interval, OR, Odds Ratio

^a^ APOE-e4 is based on the dichotomisation of APOE4 protein into carriers (≥10 NPQ) and non-carriers (<10 NPQ).

^b^ Lifestyle covariates include education, smoking status, alcohol intake, physical activity index, BMI, and presence of a self-reported health condition (one of myocardial infarction, angina, stroke, high blood pressure, high cholesterol, diabetes and cancer).

**eTable 3. Apparent and bootstrap optimism-corrected AUC for 25-year risk of Alzheimer’s disease for individual blood-based biomarkers**

| **Model** | **biomarker** | **apparent_auc^1^** | **mean_optimism^2^** | **sd_optimism^2^** | **corrected_auc^3^** | **95% CI** |
| --- | --- | --- | --- | --- | --- | --- |
| **Null^4^** | NA | 0.514 | -0.014 | 0.000 | 0.528 | (0.528, 0.528) |
| **APOE4 only^4^** | NA | 0.685 | 0.051 | 0.032 | 0.634 | (0.569, 0.699) |
| **Lifestyle only^4^** | NA | 0.622 | 0.133 | 0.032 | 0.490 | (0.425, 0.551) |
| **APOE4 + lifestyle^4^** | NA | 0.735 | 0.137 | 0.033 | 0.598 | (0.531, 0.667) |
| **Biomarker only (Model 1)^5^** | BD p-tau 217 | 0.800 | 0.098 | 0.031 | 0.703 | (0.647, 0.771) |
|  | BD p-tau 181 | 0.706 | 0.053 | 0.037 | 0.653 | (0.581, 0.724) |
|  | BD p-tau 231 | 0.658 | 0.063 | 0.039 | 0.596 | (0.519, 0.671) |
|  | Total p-tau 217 | 0.778 | 0.084 | 0.031 | 0.693 | (0.635, 0.755) |
|  | Total p-tau 231 | 0.679 | 0.049 | 0.037 | 0.630 | (0.561, 0.704) |
|  | BD p-tau 217 / Aβ42 | 0.714 | 0.083 | 0.035 | 0.631 | (0.568, 0.695) |
|  | Total p-tau 217 / Aβ42 | 0.748 | 0.086 | 0.033 | 0.662 | (0.601, 0.726) |
| **Biomarker + APOE4 (Model 2)^5^** | BD p-tau 217 | 0.816 | 0.105 | 0.028 | 0.711 | (0.659, 0.772) |
|  | BD p-tau 181 | 0.752 | 0.078 | 0.034 | 0.674 | (0.610, 0.750) |
|  | BD p-tau 231 | 0.726 | 0.084 | 0.033 | 0.642 | (0.576, 0.712) |
|  | Total p-tau 217 | 0.795 | 0.092 | 0.028 | 0.703 | (0.645, 0.759) |
|  | Total p-tau 231 | 0.737 | 0.078 | 0.032 | 0.659 | (0.596, 0.722) |
|  | BD p-tau 217 / Aβ42 | 0.758 | 0.095 | 0.030 | 0.664 | (0.610, 0.720) |
|  | Total p-tau 217 / Aβ42 | 0.781 | 0.100 | 0.028 | 0.682 | (0.631, 0.739) |
| **Biomarker + lifestyle^6^** | BD p-tau 217 | 0.831 | 0.158 | 0.027 | 0.673 | (0.622, 0.729) |
|  | BD p-tau 181 | 0.746 | 0.146 | 0.030 | 0.600 | (0.539, 0.658) |
|  | BD p-tau 231 | 0.700 | 0.146 | 0.031 | 0.553 | (0.494, 0.618) |
|  | Total p-tau 217 | 0.803 | 0.149 | 0.027 | 0.654 | (0.601, 0.705) |
|  | Total p-tau 231 | 0.707 | 0.142 | 0.030 | 0.566 | (0.505, 0.628) |
|  | BD p-tau 217 / Aβ42 | 0.738 | 0.147 | 0.030 | 0.591 | (0.537, 0.648) |
|  | Total p-tau 217 / Aβ42 | 0.769 | 0.151 | 0.029 | 0.618 | (0.567, 0.676) |
| **Biomarker + APOE4 + lifestyle^6^** | BD p-tau 217 | 0.840 | 0.158 | 0.026 | 0.683 | (0.634, 0.733) |
|  | BD p-tau 181 | 0.790 | 0.148 | 0.029 | 0.642 | (0.584, 0.700) |
|  | BD p-tau 231 | 0.763 | 0.150 | 0.030 | 0.613 | (0.554, 0.672) |
|  | Total p-tau 217 | 0.816 | 0.149 | 0.026 | 0.667 | (0.617, 0.716) |
|  | Total p-tau 231 | 0.768 | 0.146 | 0.030 | 0.622 | (0.560, 0.680) |
|  | BD p-tau 217 / Aβ42 | 0.790 | 0.153 | 0.029 | 0.637 | (0.584, 0.695) |
|  | Total p-tau 217 / Aβ42 | 0.810 | 0.156 | 0.027 | 0.654 | (0.603, 0.710) |

**^1^**: Apparent AUC is the observed AUC obtained based on the original full cohort.

**^2^**: Mean and sd of optimism over 500 bootstrap samples. For each bootstrap resampling, Optimism_b_=AUC_boot_−AUC_test_, where AUC_boot_ evaluates the model fitted based on the bootstrap sample b (b=1, …, 500), and AUC_test_ evaluates the same model as AUC_boot_ on the original data.

**^3^**: Corrected AUC: Bootstrap optimism-corrected AUC = apparent AUC – mean optimism.

**^4^**: Models without biomarkers are shown for reference in the first four rows.

**^5^**: Bootstrap optimism-corrected AUCs for Biomarker only (Model 1) and Biomarker + APOE4 (Model 2) are shown in Figure 2 in the main manuscript, and the corrected AUCs for models with lifestyle covariates are shown in supplementary Figure 2.

**eFigure 3. Association between the modifiable lifestyle risk factors and blood-based biomarkers within controls**

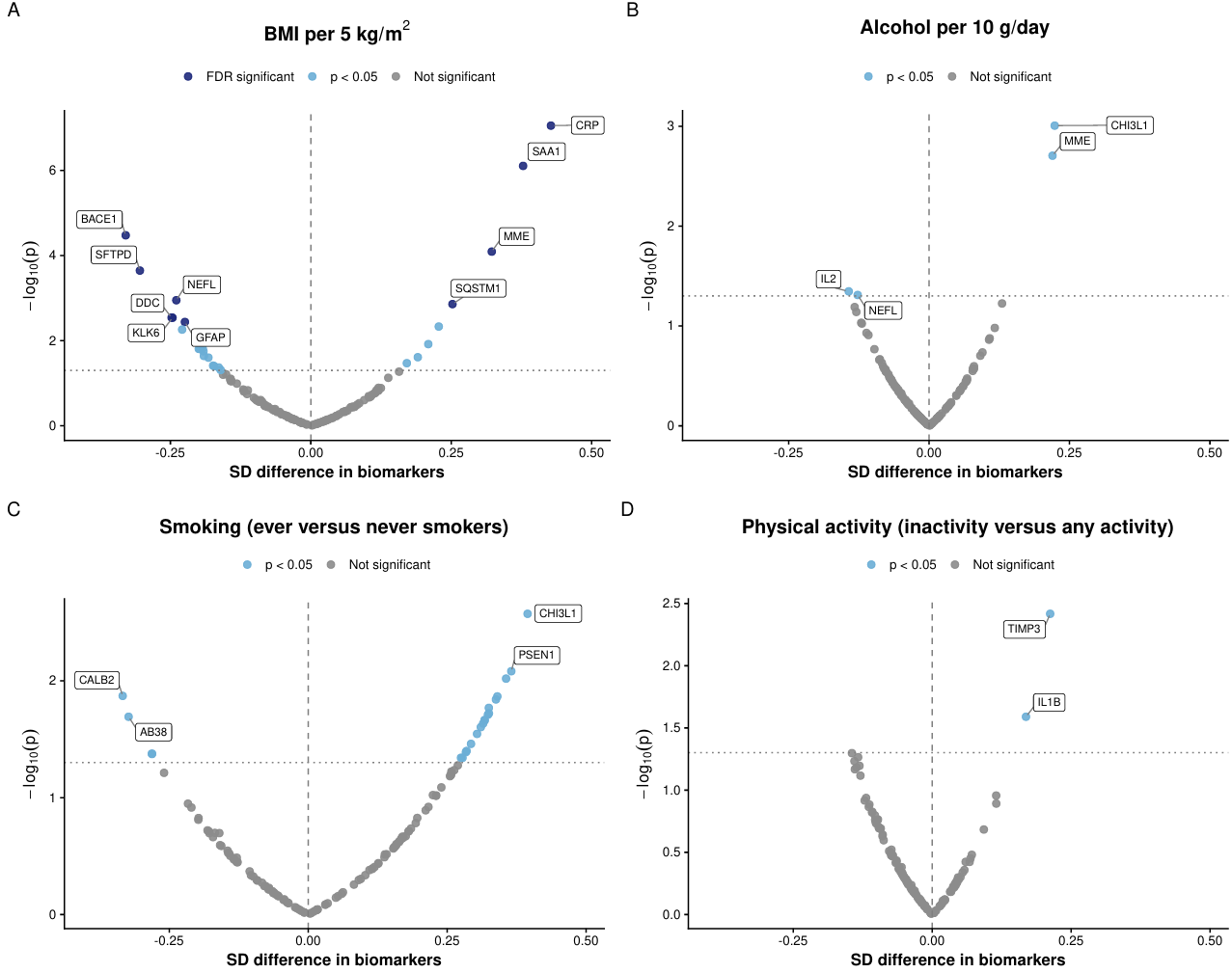

**eTable 4. Bootstrap validated incremental ΔAUC using brain-derived p-tau 217 as the base model with other blood-based biomarkers added singly**

| Biomarker added | Mean ΔAUC | Sd ΔAUC | 95% CI | | Number of replicates |
| --- | --- | --- | --- | --- | --- |
| BD p-tau 181 | 0.009 | 0.009 | (-0.001, | 0.031) | 500 |
| BD p-tau 231 | 0.021 | 0.017 | (-0.001, | 0.063) | 500 |
| Total p-tau 217 | 0.004 | 0.006 | (-0.002, | 0.021) | 500 |
| Total p-tau 231 | 0.006 | 0.008 | (-0.002, | 0.026) | 500 |
| Total p-tau 217 / Aβ42 | 0.015 | 0.012 | (-0.001, | 0.044) | 500 |
| BD p-tau 217 / Aβ42 | 0.016 | 0.013 | (-0.001, | 0.046) | 500 |

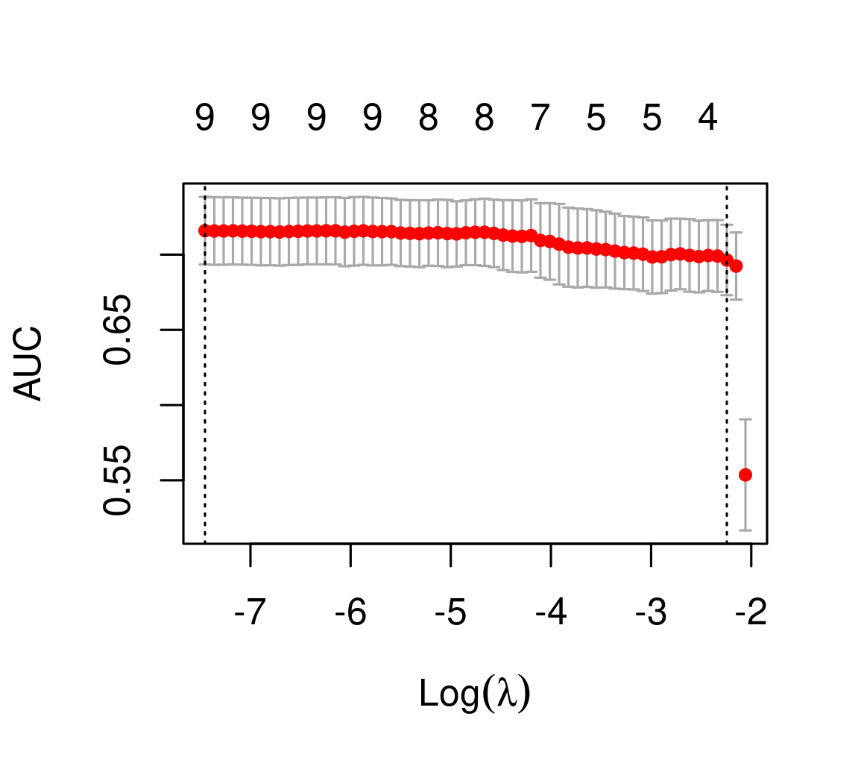
e**Figure 3. Five-fold** **cross-validated AUC versus the LASSO penalty parameter (λ)**

Notes:

- X-axis: λ is the LASSO penalty parameter. Smaller λ (weak penalty) toward the left side, and hence more variables retained).
- Y-axis: mean cross-validated AUC across the 5 folds.
- Each point represents one candidate λ value being evaluated.
- The error bars are ±1 standard error of the AUC across the cross-validation folds.
- Left vertical dashed line: lambda.min (best mean AUC).
- Right vertical dashed line: lambda.1se (largest λ whose AUC is within one standard error of the best).
- The numbers along the top are the number of non-zero predictors at each λ.

**eTable 5. LASSO model performance and selected predictors at the optimal penalty parameters**

|  | **Lambda** | **AUC^3^** | **SE** | **Number of predictors** | **Biomarkers selected**  **(penalty predictors)** | **Unpenalized predictors** |
| --- | --- | --- | --- | --- | --- | --- |
| min^1^ | 0.001 | 0.716 | 0.022 | 9 | BD p-tau 181, BD p-tau 217, BD p-tau 231,  Total p-tau 217, Total p-tau 231,  Total p-tau 217 / Aβ42 | Age,  sex,  time of blood draw |
| 1se^2^ | 0.106 | 0.696 | 0.023 | 4 | BD p-tau 217 |  |

^1^ Lambda.min: best mean cross-validated AUC.

^2^ Lambda.1se: simpler model within 1 standard error of the best.

^3^ Five-fold cross-validated AUC
